# Safety and Efficacy of Chimeric Antigen Receptor T-cell Therapy for Recurrent Glioblastoma: An Augmented Meta-analysis of Phase 1 Clinical Trials

**DOI:** 10.1101/2024.10.23.24316015

**Authors:** Ahmed Y. Azzam, Mahmoud M. Morsy, Mohammed A. Azab, Osman Elamin, Adam Elswedy, Omar S. Ahmed, Mahmoud Nassar, Ahmed Saad Al Zomia, Adham A. Mohamed, Oday Atallah, Ahmed Alamoud, Hammam A. Alotaibi, Hana J. Abukhadijah, Abdulqadir J Nashwan

## Abstract

**Introduction:** Glioblastoma is a devastating brain tumor with poor prognosis despite current treatment modalities. Chimeric antigen receptor T-cell (CAR T-cell) therapy has shown promise in other cancers but has yielded mixed results in glioblastoma. This augmented meta-analysis aims to address the limitations of previous studies and evaluate the safety and efficacy of CAR T-cell therapy for recurrent glioblastoma.

**Methods:** We followed PRISMA guidelines, including specific inclusion and exclusion criteria, for our literature review. Eight studies with 108 patients were included. We used standard and augmented meta-analyses to assess outcomes, complications, and publication bias.

**Results:** It was found that the mean overall survival for glioblastoma patients who underwent CAR T-cell therapy was 6.49 months, demonstrating no significant deviation from the median survival observed in those following the standard protocol. CAR T-cell therapy did not lead to a statistically significant improvement in achieving complete responses, with only 80% of patients exhibiting this outcome. Conversely, 44% of patients experienced stable disease, while 58% faced disease progression after CAR T-cell therapy. Adverse events were notable, with CAR T cell therapy-related encephalopathy affecting 37% of treated patients, while cytokine release syndrome was a rare event, observed in only 3% of cases.

**Conclusions:** To our knowledge, this is the first study that utilizes this novel statistical technique to predict the outcomes of CAR T-cell therapy for recurrent glioblastoma. The results of this study are predictive rather than confirmatory. CAR T-cell therapy for glioblastoma was not predicted to significantly improve survival or achieve substantial complete responses. Stable disease rates are modest, while disease progression is notable. Adverse events, especially CAR T-cell therapy-related encephalopathy, raise safety concerns. Further trials and refinements are needed to enhance CAR T-cell therapy’s effectiveness and safety in glioblastoma treatment, Manuscript Click here to view linked References potentially through optimizing administration routes and target antigens or combining it with other therapies. This challenging disease necessitates continued research to improve patient outcomes.

## 1. Introduction

Glioblastoma is the most common and the most aggressive primary brain tumor in adults with a poor prognosis. The current standard treatment protocol consists of surgical resection of the tumor, temozolomide as chemotherapy, followed by adjuvant radiotherapy. Despite the current advances in oncology, the prognosis of glioblastoma remains poor, with median overall survival ranging between 16 to 18 months in clinical trials [1–3].

Immunotherapy is one of the most important breakthroughs in cancer treatment and has been reported to have favorable survival rates and a varying safety profile. Chimeric antigen receptor T cell (CAR T-cell) is a genetically modified type of lymphocyte synthesized through advanced biomedical engineering techniques to target specific tumor antigens with the aim of eliminating them. CAR T-cell lymphocytes have shown promising results in hematological malignancies. In contrast, the results for solid tumors were not satisfactory and had limited efficacy in improving survival outcomes [4–7].

The safety and efficacy of CAR T-cell therapy for glioblastoma have been investigated in several clinical trials in the literature, showing a highly variable response rate that ranges from low to high response rates. In the current literature, only one meta-analysis has explored CAR T-cell therapy for glioblastoma. However, this meta-analysis had several limitations, including a limited number of included studies and a high risk of bias due to variations in results and data sparsity [8, 9].

Based on these limitations, we conducted an augmented meta-analysis to examine the safety and efficacy of CAR T-cell therapy for recurrent glioblastoma. The primary objective of this augmented meta-analysis is to address the shortcomings of the study by Jang JK et al. [10] by increasing the number of included studies through the synthetic generation of new data points using a novel augmentation technique. This approach allows us to overcome the limited number of studies, reduce the risk of bias, and alleviate data sparsity.

What makes this meta-analysis unique is the combination of both a standard meta-analysis and the use of this novel statistical technique. Our analysis will employ two different statistical methods: the first will be a standard meta-analysis, while the second analysis will utilize this innovative statistical technique to augment the number of included studies by randomly calculating averages from the original data, resulting in new synthetic data. This approach aims to mitigate the limitations associated with low sample size and the risk of bias due to the limited number of studies.

## 2. Methods

### 2.1. Methodology and Inclusion Criteria

This study was conducted in accordance with the Preferred Reporting Items for Systematic Reviews and Meta-Analysis (PRISMA) guidelines [11]. The inclusion criteria for our meta-analysis included: patients who underwent CAR-T cell therapy for glioblastoma, original articles, and English-language articles. Articles which did not investigate the safety and efficacy of CAR-T cell therapy for glioblastoma, review articles, systematic reviews, meta-analysis, case reports, case series, editorial letters, conference abstracts, and non-English articles were excluded.

### 2.2. Literature Review and Risk of Bias Assessment

We searched PubMed/Medline, EMBASE, Scopus, Web of Science and Google Scholar databases with the relevant search strategy and keywords up to 29^th^ of September 2023. The search terms included “Glioblastoma” AND “CAR-T Cell Therapy” AND “Chimeric Antigen Receptor” AND “High-grade Glioma” AND “Immunotherapy” AND “Neuro-Oncology”.

Study quality was assessed using the Newcastle Ottawa Scale (NOS). Each study was evaluated in three categories: the proper selection of the study population, the comparability of the study groups, and 3) the ascertainment of the exposure or outcome of interest. Studies with a score of seven or more (out of a maximum of nine points) were considered high-quality studies. Studies that were ambiguous or led to differences in opinion between the two reviewers were re-evaluated during a consensus meeting that included a third reviewer. Three authors independently evaluated the risk of bias. The reviewers settled the discrepancies by discussion.

### 2.3. Statistical Analysis

We performed a meta-analysis of the included studies to estimate the cumulative incidence (event rate) and 95% confidence interval (CI), continuous outcomes were estimated through mean of raw data (MRAW) and 95% confidence interval (CI). The heterogeneity of results among the included studies was examined using Cochrane’s Q-test and the I^2^ statistic. The common-effects (fixed-effect) model was indicated for outcomes without significant heterogeneity, while random-effects model was indicated for outcomes with significant heterogeneity. Heterogeneity was assessed through visual inspection of the forest plots and measured using the I^2^ and chi-square ( 2) tests. The 2 test was employed to determine the presence of significant heterogeneity, while the I^2^ test was utilized to quantify the magnitude of heterogeneity, if present. The interpretation of the I^2^ test followed the recommendations provided by the Cochrane Handbook (Part 2, Chapter 9). For testing statistical heterogeneity, a significance level (α) below 0.1 was considered indicative of significant heterogeneity, as recommended by the Cochrane Handbook. Publication bias was visually assessed with a funnel plot and confirmed by Egger’s test. In cases of publication bias, a trim-and-fill adjustment was performed to calculate the adjusted pooled proportion and MRAW. All p-values were two-sided, and a p-value < 0.05 was considered statistically significant. Also, statistical significance was assessed in alliance with confidence interval range. The analysis was conducted using the R version 4.3.0. In addition, an individual patient meta-analysis was conducted to assess predictors of outcomes and complications.

### 2.4. Augmented Meta-analysis

To increase the statistical power and precision of our meta-analysis, we utilized an augmented meta-analysis technique. This involves artificially amplifying the number of included studies by calculating the averaged values between randomly selected pairs of studies from the original dataset. For example, if our meta-analysis originally included six studies, we would generate six additional artificial studies by randomly selecting two of the original studies and averaging their results. This results in 12 studies total in the augmented meta-analysis (the original six studies plus six artificially generated averages). By increasing the number of studies, we can improve the stability and reliability of the meta-analytic estimates. However, it is important to note that the artificially generated averaged studies do not provide fully independent data points. The augmented meta-analysis aims to strike a balance between maximizing information and minimizing dependence among the synthesized studies.

## 3. Results

### 3.1. Literature Review and Risk of Bias Assessment

Eligible articles screening, inclusion and exclusion process was done following PRISMA guidelines as shown in **Figure 1**. The risk of bias assessment according to the NOS scale is listed in **Table 1**.

**Figure 1:**
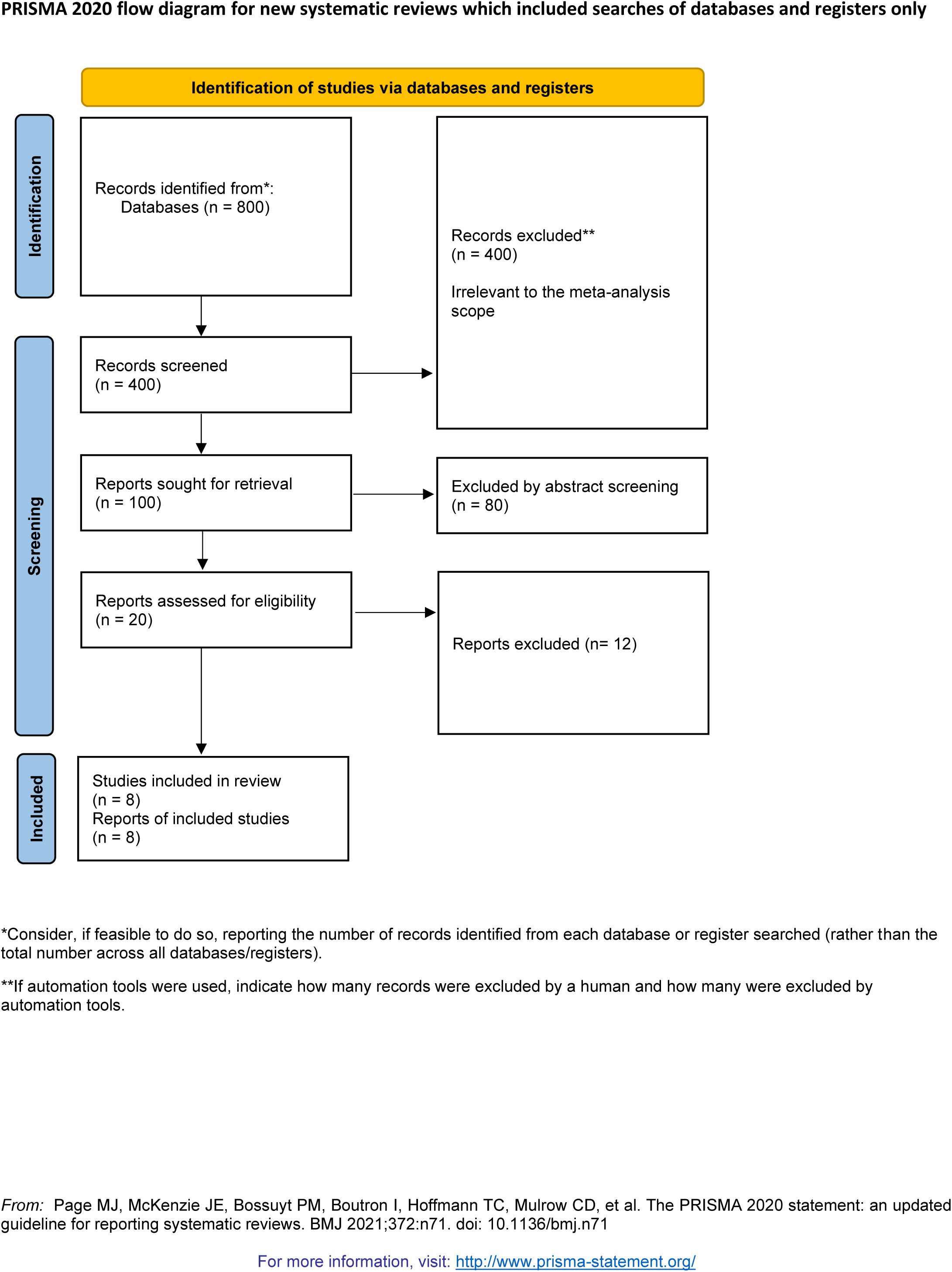
PRISMA Chart Flow for the Included Studies.

**Table 1:**
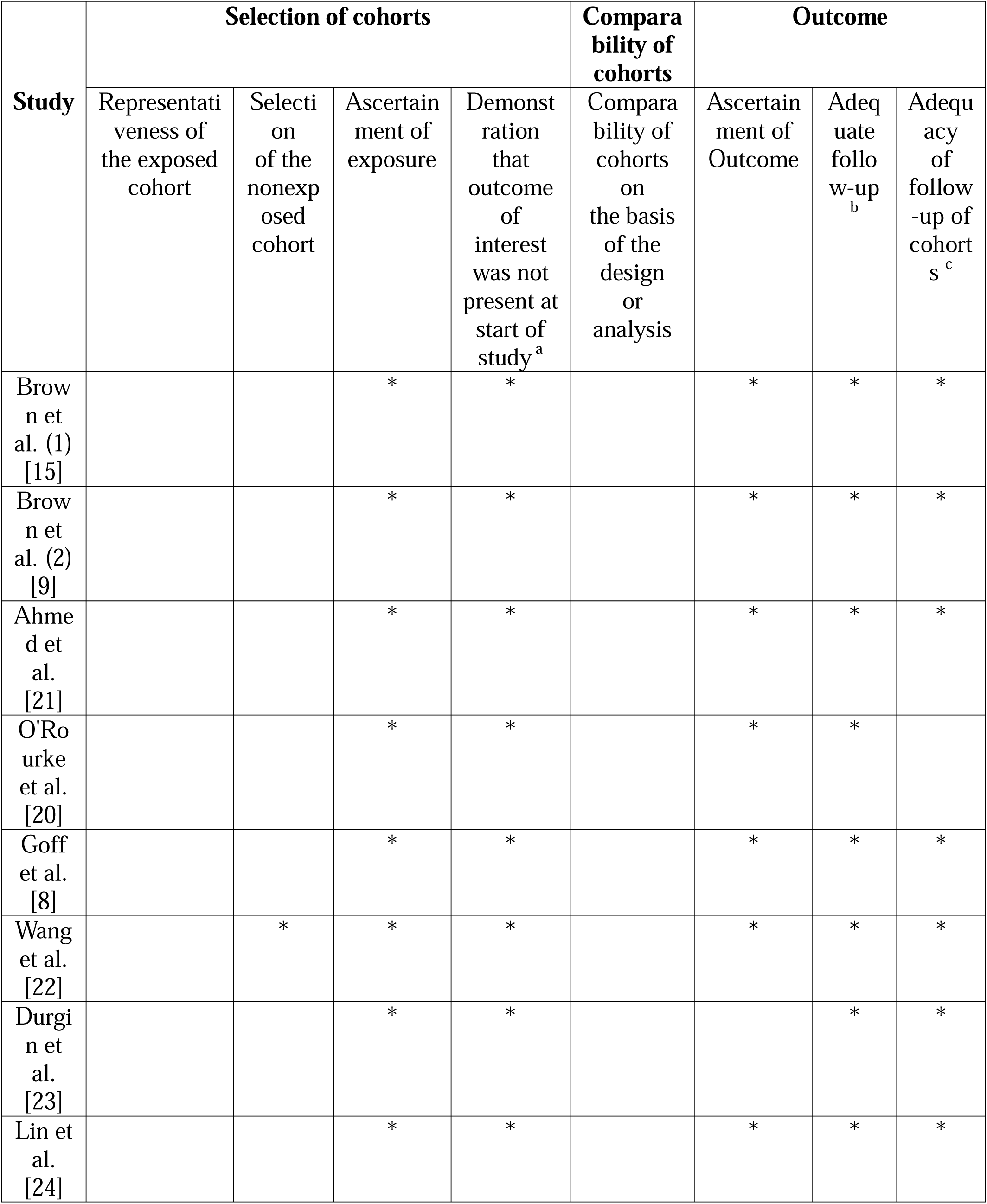
Newcastle-ottawa scale (NOS) risk of bias assessment for the included studies.

### 3.2. Included Studies Characteristics

We have included eight studies with a total of 108 patients (**Table 2**). Two studies aimed to target IL13Ra2 as a therapeutic target, four studies aimed to target EGFRvIII, one study focused on HER2, and one study centered around EphA2. Only two studies used intracranial access as a delivery method, while the other studies utilized intravenous access. The maximum tolerated dose in the included studies was 1 x 10^8^ m^2^ (**Table 3**). Information regarding overall survival after CAR T-cell therapy administration, outcomes, and adverse events is listed in **Table 4**.

**Table 2:** Baseline Characteristics of the Included Clinical Trials.

| Study | Clinical Trial Phase | Year | Number of Patients | Standard Treatment Protocol | IDH-Mutation Status |  | MGMT-Mutation Status |  |
| --- | --- | --- | --- | --- | --- | --- | --- | --- |
|  |  |  |  |  | Positive | Negative | Positive | Negative |
| Brown et al. (1) [15] | N/A | 2015 | 3 | N/A | N/A | N/A | N/A | N/A |
| Brown et al. (2) [9] | Phase 1 | 2016 | 1 | Surgical Resection, Chemotherapy, and Temozolomide | 1 | 0 | 0 | 1 |
| Ahmed et al. [21] | Phase 1 | 2017 | 36 | Surgical Resection, Chemotherapy, and Temozolomide | N/A | N/A | N/A | N/A |
| O'Rourke et al. [20] | Phase 1 | 2017 | 20 | Surgical Resection, Chemotherapy, and Temozolomide | N/A | N/A | 0 | 1 |
| Goff et al. [8] | Phase 1 | 2019 | 22 | Surgical Resection, Chemotherapy, and Temozolomide | N/A | N/A | 4 | 14 |
| Wang et al. [22] | N/A | 2019 | 20 | Surgical Resection, Chemotherapy, and Temozolomide | N/A | N/A | N/A | N/A |
| Durgin et al. [23] | N/A | 2021 | 1 | Surgical Resection and Chemotherapy | 1 | 0 | 1 | 0 |
| Lin et al. [24] | Phase 1 | 2021 | 5 | Surgical Resection, Chemotherapy, and Temozolomide | N/A | N/A | N/A | N/A |

**Table 3:** Therapeutic Characteristics of the Included Trials.

| Study | Therapeutic Target | Delivery Method | T-cell dose, | T-cell Persistence | Maximum Tolerated |
| --- | --- | --- | --- | --- | --- |

|  |  |  | **m<sup>2</sup>** |  | **Dose, m<sup>2</sup>** |
| --- | --- | --- | --- | --- | --- |
| Brown et al.<br>(1) [15] | IL13Ra2 | Intracranial | (9.6 x 10<sup>8</sup>) – (10.6 x 10<sup>8</sup>) | N/A | 1 x 10<sup>8</sup> |
| Brown et al.<br>(2) [9] | IL13Ra2 | Intracranial | (2 x 10<sup>6</sup>) – (10 x 10<sup>6</sup>) | More than one week | N/A |
| Ahmed et al. [21] | HER2 | Intravenous | (1 x 10<sup>6</sup>) – (1 x 10<sup>8</sup>) | More than one year | 1 x 10<sup>8</sup> |
| O'Rourke et al. [20] | EGFRvIII | Intravenous | (1 x 10<sup>8</sup>) – (5 x 10<sup>8</sup>) | More than one month | 1 x 10<sup>8</sup> |
| Goff et al. [8] | EGFRvIII | Intravenous | (1 x 10<sup>7</sup>) – (6 x 10<sup>10</sup>) | More than three-month | 1 x 10<sup>8</sup> |
| Wang et al. [22] | EGFRvIII | N/A | N/A | N/A | N/A |
| Durgin et al. [23] | EGFRvIII | Intravenous | 9.2 x 10<sup>7</sup> | More than three-years | N/A |
| Lin et al. [24] | EphA2 | Intravenous | 1 x 10<sup>6</sup> ** | More than one month | N/A |

\*\* The Unit: Cells/KG

**Table 4:** Survival Outcomes and Adverse Events.

| **Study** | **Overall Survival , Median Months** | **Total Patients, n** | **Complete Response, n** | **Stable Disease, n** | **Progressive Disease, n** | **Cytokine Release Syndrome, n** | **CAR-T Cell Related Encephalopathy, n** |
| --- | --- | --- | --- | --- | --- | --- | --- |

|  | (IQR) |  |  |  |  |  |  |
| --- | --- | --- | --- | --- | --- | --- | --- |
| Brown et al. (1) [15] | 10.3 (8.6 - 13.9) | 3 | 2 | 0 | 1 | 0 | 2 |
| Brown et al. (2) [9] | 9.8 | 1 | 1 | 0 | 0 | 1 | 1 |
| Ahmed et al. [21] | 11.1 (4.1 - 27.2) | 36 | 0 | 7 | 8 | 0 | 2 |
| O'Rourke et al. [20] | 8.3 (3.3 - 14.8) | 20 | 0 | 5 | 1 | 0 | 1 |
| Goff et al. [8] | 6.9 (2.8 - 10) | 22 | 0 | 0 | 17 | 2 | 10 |
| Wang et al. [22] | 8.1 (3.4 - 9.5) | 20 | 0 | 3 | 7 | N/A | N/A |
| Durgin et al. [23] | 34 | 1 | N/A | N/A | N/A | 1 | N/A |
| Lin et al. [24] | 5.4 (2.8 - 5.9) | 5 | 0 | 1 | 2 | 2 | 0 |

### 3.3. Standard Meta-analysis Results

#### 3.3.1. Overall Survival

The mean overall survival for patients who received CAR T-cell therapy was **7.01 months, with a 95% CI= (5.23 – 8.78),** (**Figure 2**). No significant heterogeneity was observed (I^2^ = 0%), so the common-effects model was used. Visual inspection of the funnel plot for asymmetry (**Supplementary Figure 1**) did not suggest a risk of publication bias. Egger’s test for publication bias could not be performed since the number of included studies in this outcome was less than ten.

**Figure 2:**
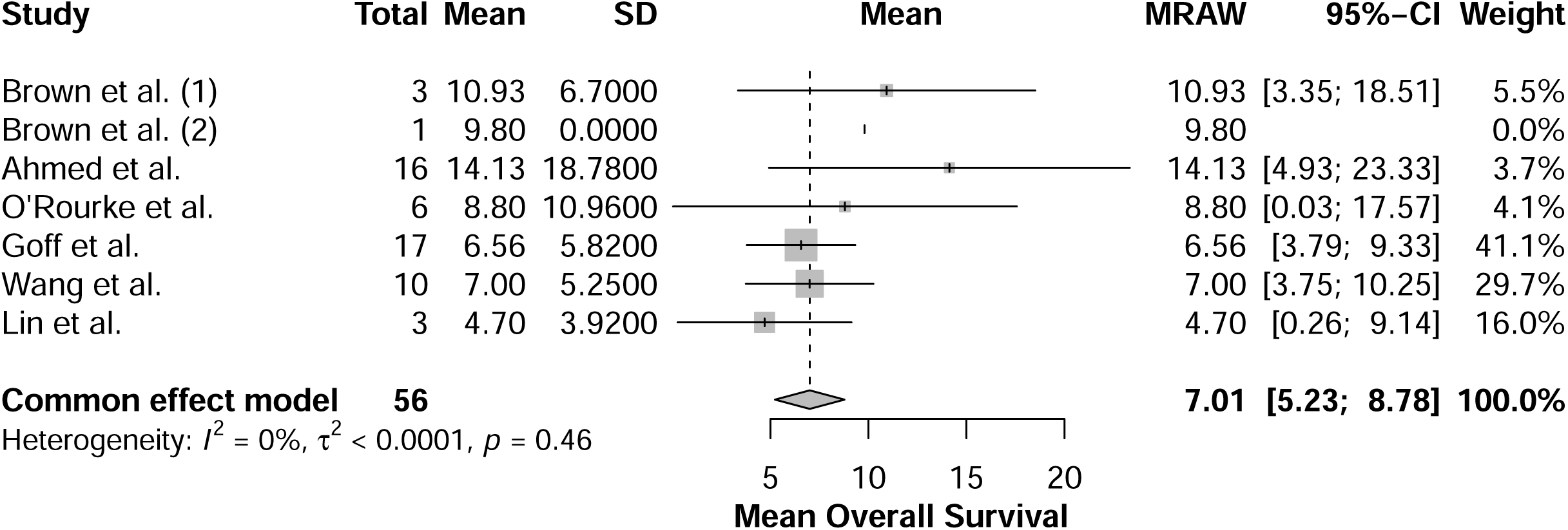
Forest plot for mean overall survival.

#### 3.3.2. Complete Response

The proportion of patients who achieved a complete response for glioblastoma after receiving CAR T-cell therapy was **80%, with a 95% CI= (17% – 100%)** (**Figure 3**). However, the results were not suggestive of statistical significance, since the confidence interval range has crossed the value of 1.0, 95% CI= (0.17 – 1.00). No significant heterogeneity was observed (I^2^ = 0%), so the common-effects model was used. The wide range of the 95% confidence interval was suggestive of data sparsity, so augmented meta-analysis was indicated before making conclusions.

**Figure 3:**
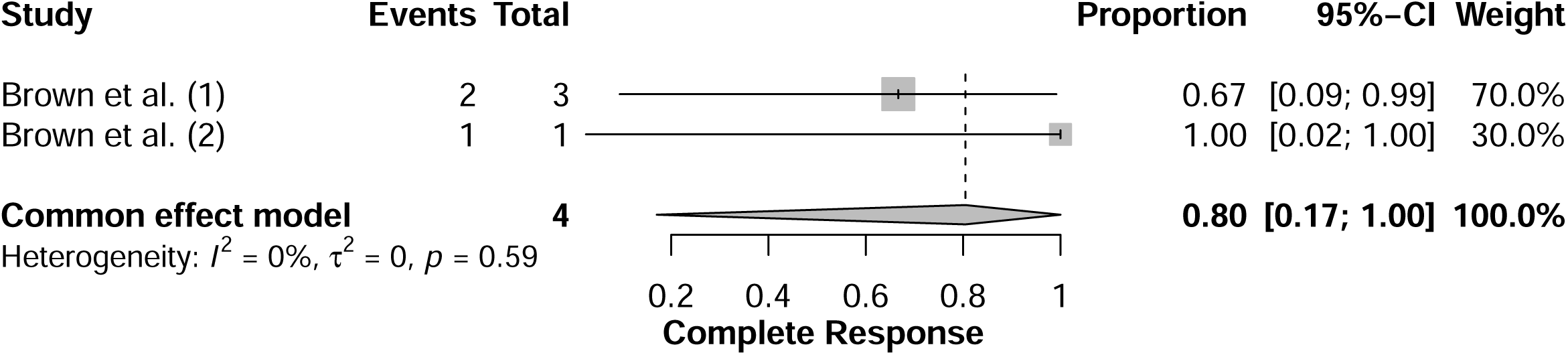
Forest plot for complete response outcome.

Visual inspection of the funnel plot for asymmetry (**Supplementary Figure 2**) was not possible since there were only two studies included in this outcome analysis. Additionally, Egger’s test for publication bias could not be performed since the number of included studies in this outcome was less than ten.

#### 3.3.3. Stable Disease

The proportion of patients who maintained a stable disease condition after receiving CAR T-cell therapy was **46%, with a 95% CI= (28% – 64%),** (**Figure 4**). No significant heterogeneity was observed (I^2^ = 29%, p-value= 0.24), so the common-effects model was used. The wide range of the 95% confidence interval was suggestive of data sparsity, so augmented meta-analysis was indicated before making conclusions. Visual inspection of the funnel plot for asymmetry (**Supplementary Figure 3**) did not suggest a risk of publication bias. Egger’s test for publication bias could not be performed since the number of included studies in this outcome was fewer than ten.

**Figure 4:**
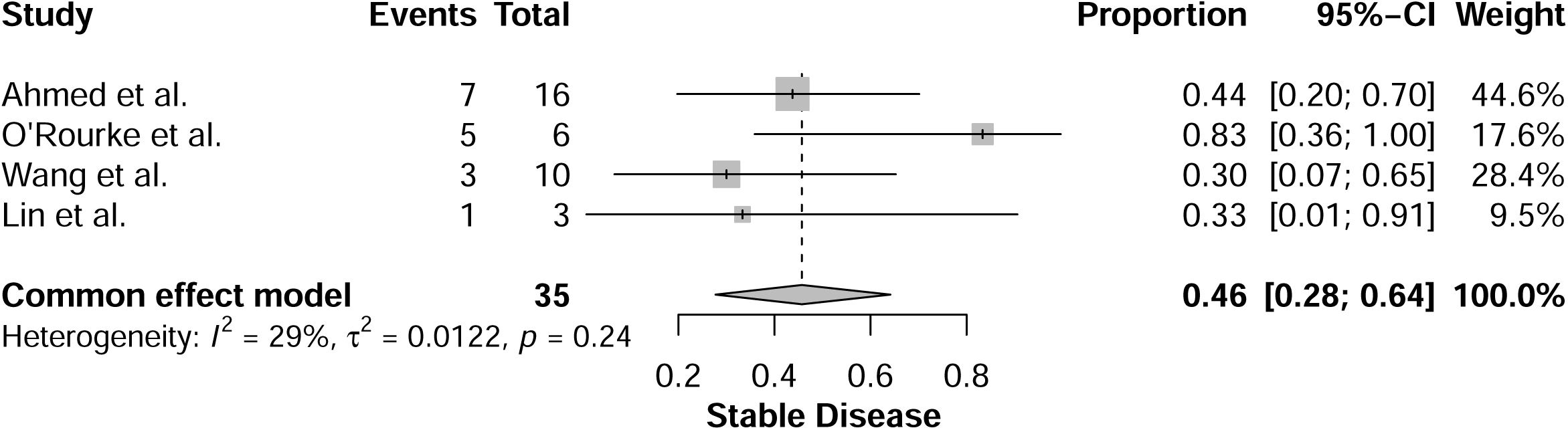
Forest plot for stable disease outcome.

#### 3.3.4. Disease Progression

The proportion of patients who went into disease progression and worsening of their condition after receiving CAR T-cell therapy was **63%, with a 95% CI= (27% – 93%),** (**Figure 5**).

**Figure 5:**
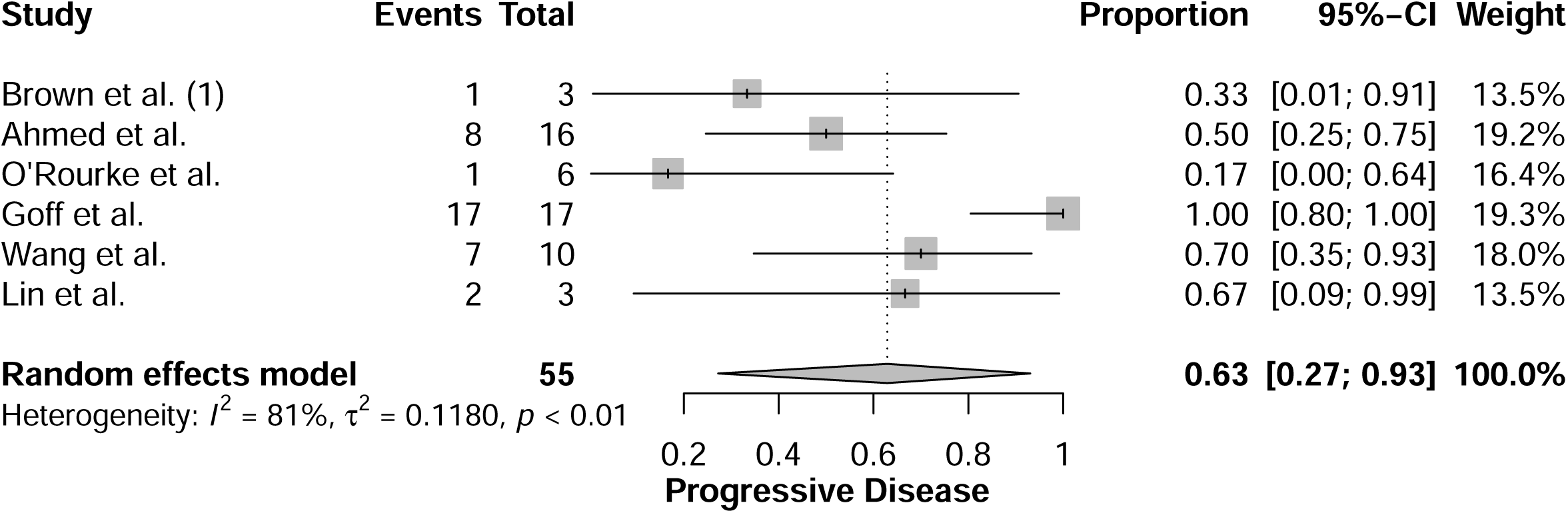
Forest plot for progressive disease outcome.

Significant heterogeneity was observed (I^2^ = 81%, p-value < 0.01), so the random-effects model was used. Sensitivity analysis was conducted to resolve the significant heterogeneity, after excluding Goff et al. study the heterogeneity was resolved (I^2^= 10%, p-value= 0.35) resulting a proportion of **50%, with a 95% CI= (31% – 68%)**. The wide range of the 95% confidence interval was suggestive of data sparsity, so augmented meta-analysis was indicated before making conclusions. Visual inspection of the funnel plot for asymmetry (**Supplementary Figure 4**) did not suggest a risk of publication bias. Egger’s test for publication bias could not be performed since the number of included studies in this outcome was fewer than ten.

#### 3.3.5. CAR T-Cell Therapy-Related Encephalopathy

The proportion of patients who developed CAR T-cell therapy related encephalopathy as an adverse event from CAR T-cell therapy was **31%, with a 95% CI= (5% – 64%),** (**Figure 6**). Significant heterogeneity was observed (I^2^ = 62%, p-value < 0.01), so the random-effects model was used. Sensitivity analysis was conducted to address the significant heterogeneity; however, the heterogeneity remained unresolved (I^2^ = 68%, p-value = 0.02). This raise concerns that the source of heterogeneity may not be attributed to anomalous results, but rather to variations in measurements across the included studies regarding this adverse event. We attempted to investigate the reasons behind this statistically significant heterogeneity; however, we were uncertain about its cause. We hypothesized that the most likely explanation could be the lack of details provided in the included studies, which may indicate a potential reporting bias for this adverse event. The wide range of the 95% confidence interval was suggestive of data sparsity, so augmented meta-analysis was indicated before making conclusions. Visual inspection of the funnel plot for asymmetry (**Supplementary Figure 5**) did not suggest a risk of publication bias. Egger’s test for publication bias could not be performed since the number of included studies in this outcome was fewer than ten.

**Figure 6:**
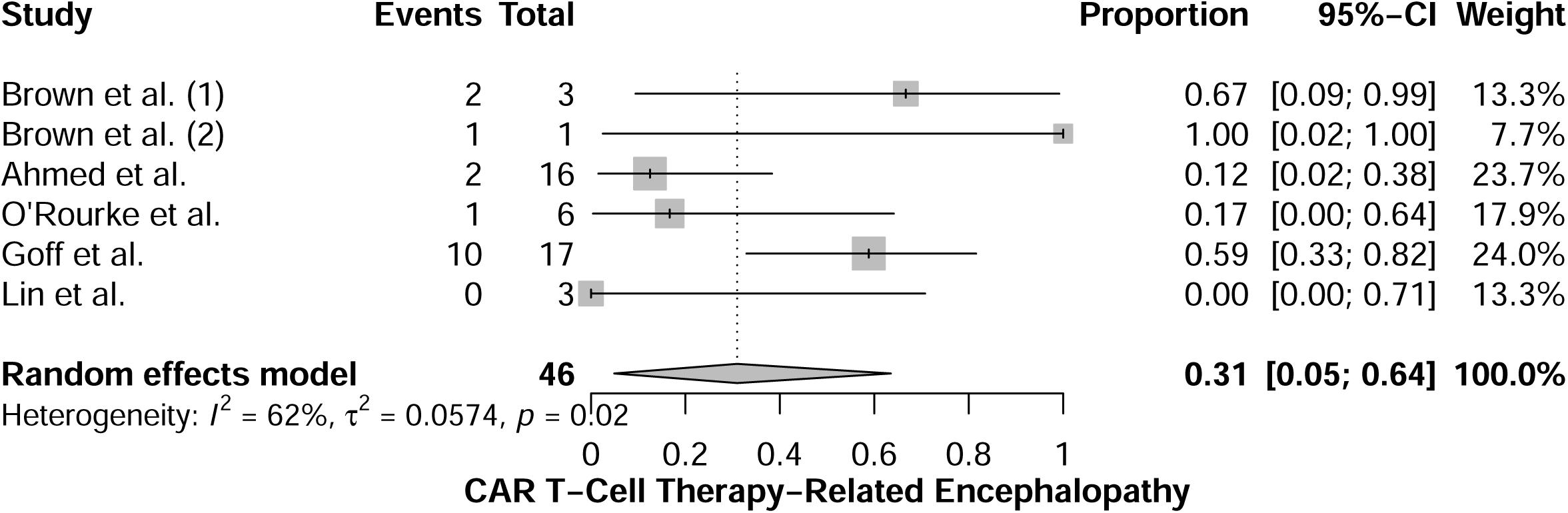
Forest plot for CAR T-cell Therapy-Related Encephalopathy adverse event.

#### 3.3.6. Cytokine Release Syndrome

The proportion of patients who developed cytokine release syndrome as an adverse event from CAR T-cell therapy was **2%, with a 95% CI= (0% – 13%),** (**Figure 7**). Significant heterogeneity was observed (I^2^ = 65%, p-value < 0.01), so the random-effects model was used. Sensitivity analysis was conducted to resolve the significant heterogeneity, after excluding Brown et al. (2), Durgin et al., and Lin et al. studies the heterogeneity was resolved (I^2^= 0%, p-value= 0.5). The proportion after resolving the heterogeneity was **0%, with a 95% CI= (0% – 4%)**. The wide range of the 95% confidence interval was suggestive of data sparsity, so augmented meta-analysis was indicated before making conclusions. Visual inspection of the funnel plot for asymmetry (**Supplementary Figure 6**) did not suggest a risk of publication bias. Egger’s test for publication bias could not be performed since the number of included studies in this outcome was fewer than ten.

**Figure 7:**
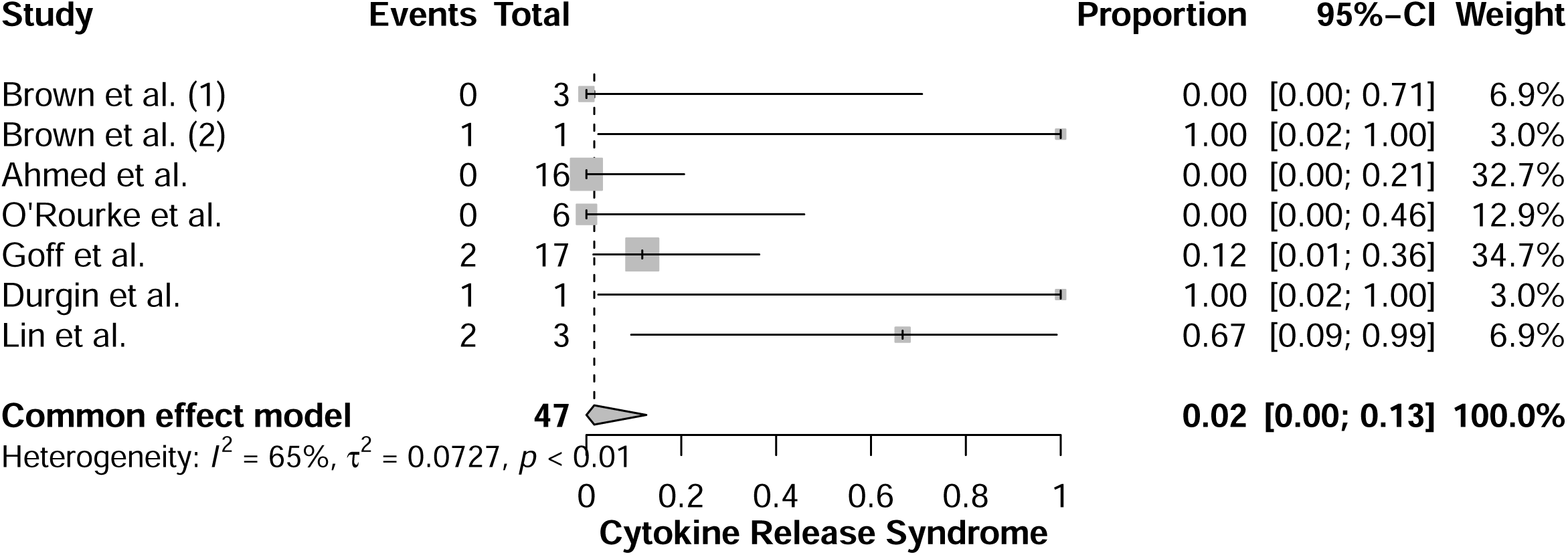
Forest plot for cytokine release syndrome adverse event.

### 3.4. Augmented Meta-analysis

#### 3.4.1. Overall Survival

After conducting an augmented analysis, the mean overall survival for patients who received CAR T-cell therapy was **6.94 months, with a 95% CI= (5.73 – 8.15)** (**Figure 8**). No significant heterogeneity was observed (I^2^ = 0%, p-value= 0.88), so the common-effects model was used.

**Figure 8:**
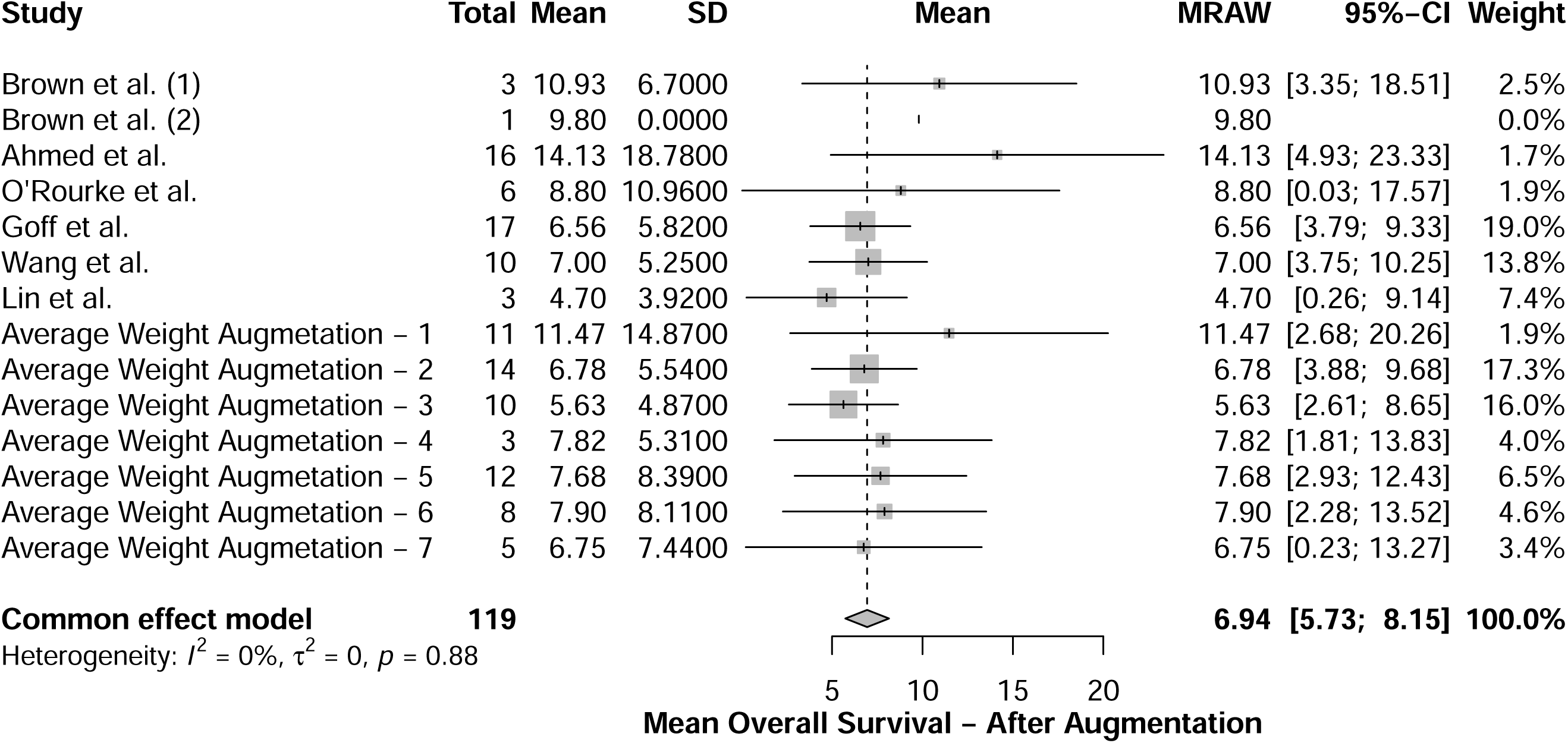
Forest plot for mean overall survival after augmentation.

Upon visual inspection of the funnel plot for asymmetry (**Supplementary Figure 7**), a potential risk of publication bias was identified. Therefore, Egger’s regression test was employed to confirm the presence of publication bias. The results of Egger’s regression test (t = 3.55, df = 11, p-value = 0.0046) indicated a significant risk of publication bias, prompting the use of the trim– and-fill technique. The mean overall survival, following the application of the trim-and-fill method for the augmented analysis, was **6.49 months, with a 95% CI= (5.36 – 7.63)**. This represents a decrease of 0.52 months in overall survival when compared to the standard analysis prior to augmentation.

#### 3.4.2. Complete Response

After conducting an augmented analysis, the proportion of patients who developed complete response of the disease after receiving CAR T-cell therapy was **70%, with a 95% CI= (19% – 100%)** (**Figure 9**). No significant heterogeneity was observed (I^2^ = 0%, p-value= 0.75), so the common-effects model was used. Visual inspection of the funnel plot for asymmetry (**Supplementary Figure 8**) did not suggest a risk of publication bias. Egger’s test for publication bias could not be performed since the number of included studies in this outcome was fewer than ten. However, the results were not statistically significant compared to the analysis conducted prior to augmentation. This suggests that the current evidence may not provide sufficient evidence to determine the efficacy of CAR T-cell therapy in achieving a complete response for glioblastoma patients.

**Figure 9:**
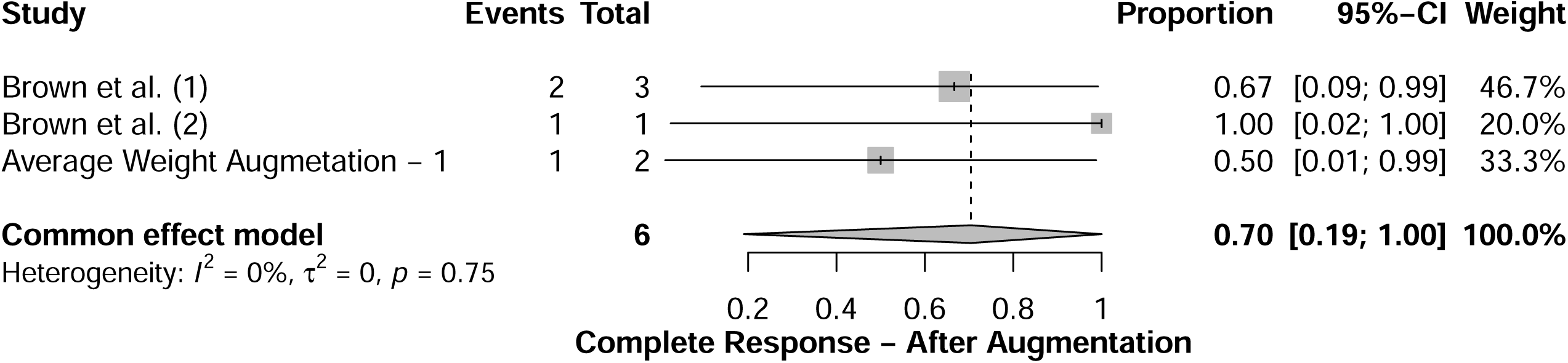
Forest plot for complete response outcome after augmentation.

#### 3.4.3. Stable Disease

After conducting an augmented analysis, the proportion of patients who had stable disease who received CAR T-cell therapy was **44%, with a 95% CI= (31% – 58%),** (**Figure 10**). No significant heterogeneity was observed (I^2^ = 0%, p-value= 0.61), so the common-effects model was used. Visual inspection of the funnel plot for asymmetry (**Supplementary Figure 9**) did not suggest a risk of publication bias. Egger’s test for publication bias could not be performed since the number of included studies in this outcome was fewer than ten. This signifies a 2% decrease in patients who maintained stable disease after receiving therapy, compared to the analysis conducted before augmentation.

**Figure 10:**
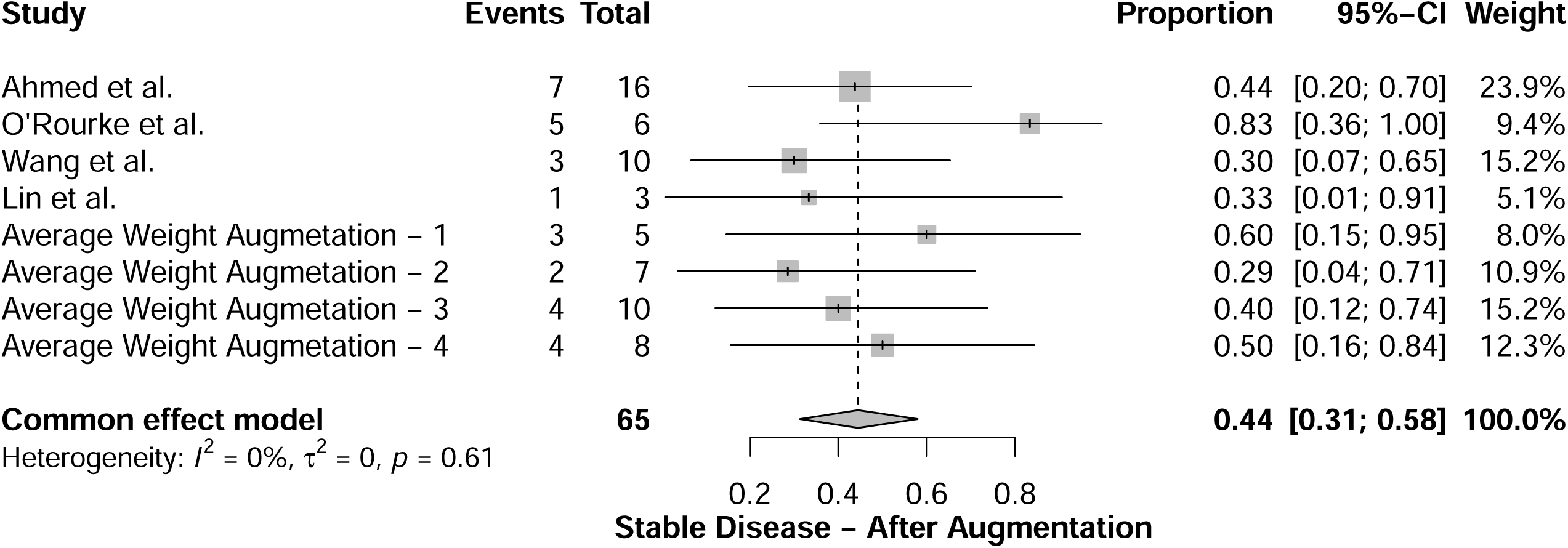
Forest plot for stable disease outcome after augmentation.

#### 3.4.4. Disease Progression

After conducting an augmented analysis, the proportion of patients who had progression in their disease after receiving CAR T-cell therapy was **62%, with a 95% CI= (45% – 79%),** (**Figure 11**). Significant heterogeneity was observed (I^2^ = 64%, p-value < 0.01), so the random-effects model was used. Sensitivity analysis was used to resolve the significant heterogeneity, the heterogeneity was resolved (I^2^= 0%, p-value= 0.6), after excluding Goff et al. study, and the new proportion after resolving heterogeneity is **58%, with a 95% CI= (47% – 68%)**.

**Figure 11:**
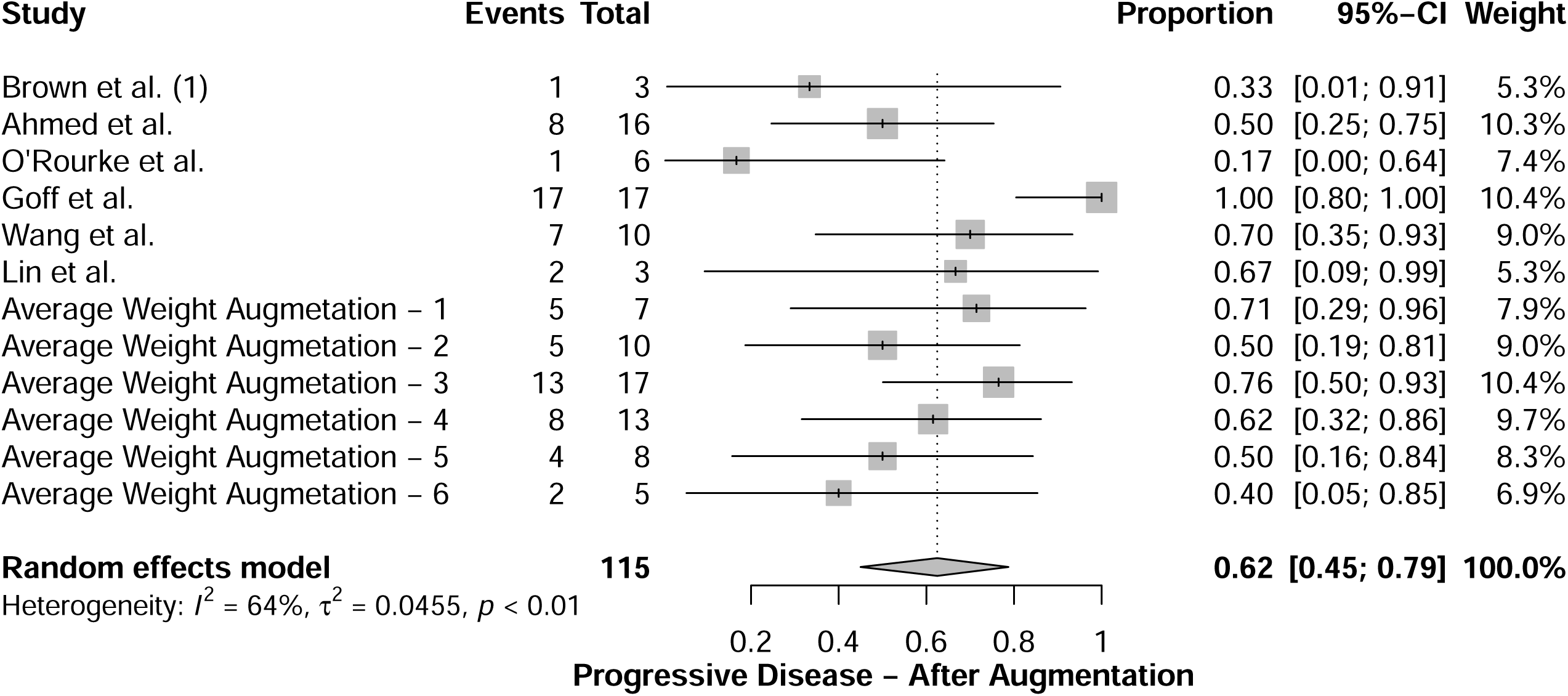
Forest plot for progressive disease outcome after augmentation.

Visual inspection of the funnel plot for asymmetry (**Supplementary Figure 10**) did not suggest a risk of publication bias. Egger’s regression test was employed to confirm the presence of publication bias. The results of Egger’s regression test (t = –1.52, df = 9, p-value = 0.1640) clarified that there was no significant risk of publication bias. This signifies an 8% increase in patients who had disease progression after receiving CAR T-cell therapy, compared to the analysis conducted before augmentation.

#### 3.4.5. CAR T-Cell Therapy-Related Encephalopathy

The proportion of patients who developed CAR T-cell therapy-related encephalopathy as an adverse event after conducting the augmented analysis was **36%, with a 95% CI= (21% – 53%)** (**Figure 12**). Significant heterogeneity was observed (I^2^ = 39%, p-value= 0.08), so the random-effects model was used. Sensitivity analysis was conducted to overcome the significant heterogeneity, after excluding Brown et al. (2) and Lin et al. studies the heterogeneity was resolved (I^2^= 36%, p-value= 0.12) resulting in a proportion of **37%, with a 95% CI= (22% – 53).** Visual inspection of the funnel plot for asymmetry (**Supplementary Figure 11**) did not suggest a risk of publication bias. Egger’s regression test was employed to confirm the presence of publication bias. The results of Egger’s regression test (t = 0.43, df = 8, p-value = 0.6812) clarified that there was no significant risk of publication bias. This signifies a 6% increase in the incidence of developing CAR T-cell therapy-related encephalopathy as an adverse event after receiving therapy, compared to the analysis conducted before augmentation.

**Figure 12:**
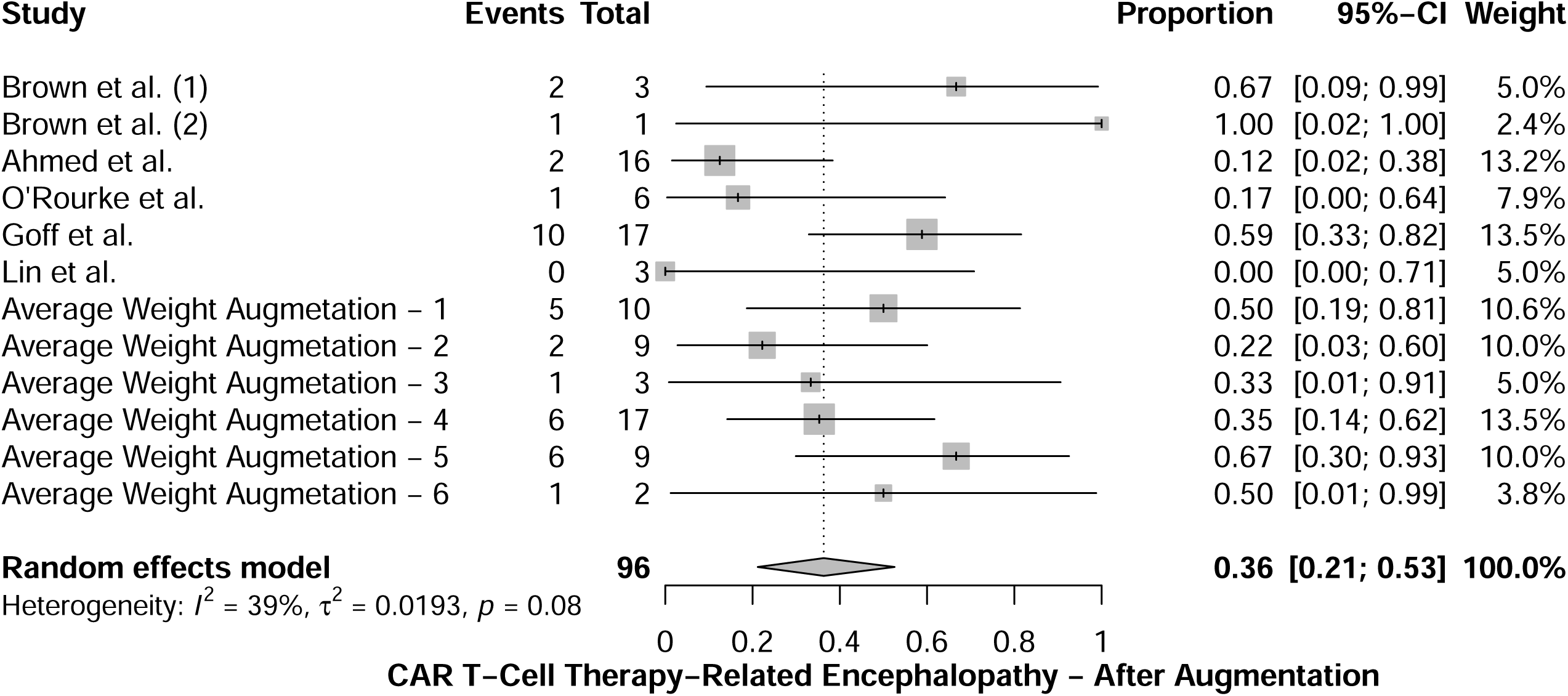
Forest plot for CAR T-cell Therapy-Related Encephalopathy adverse event after augmentation.

#### 3.4.6. Cytokine Release Syndrome

The proportion of patients who developed cytokine release syndrome as an adverse event after conducting the augmented analysis was **12%, with a 95% CI= (1% – 29%)** (**Figure 13**).

**Figure 13:**
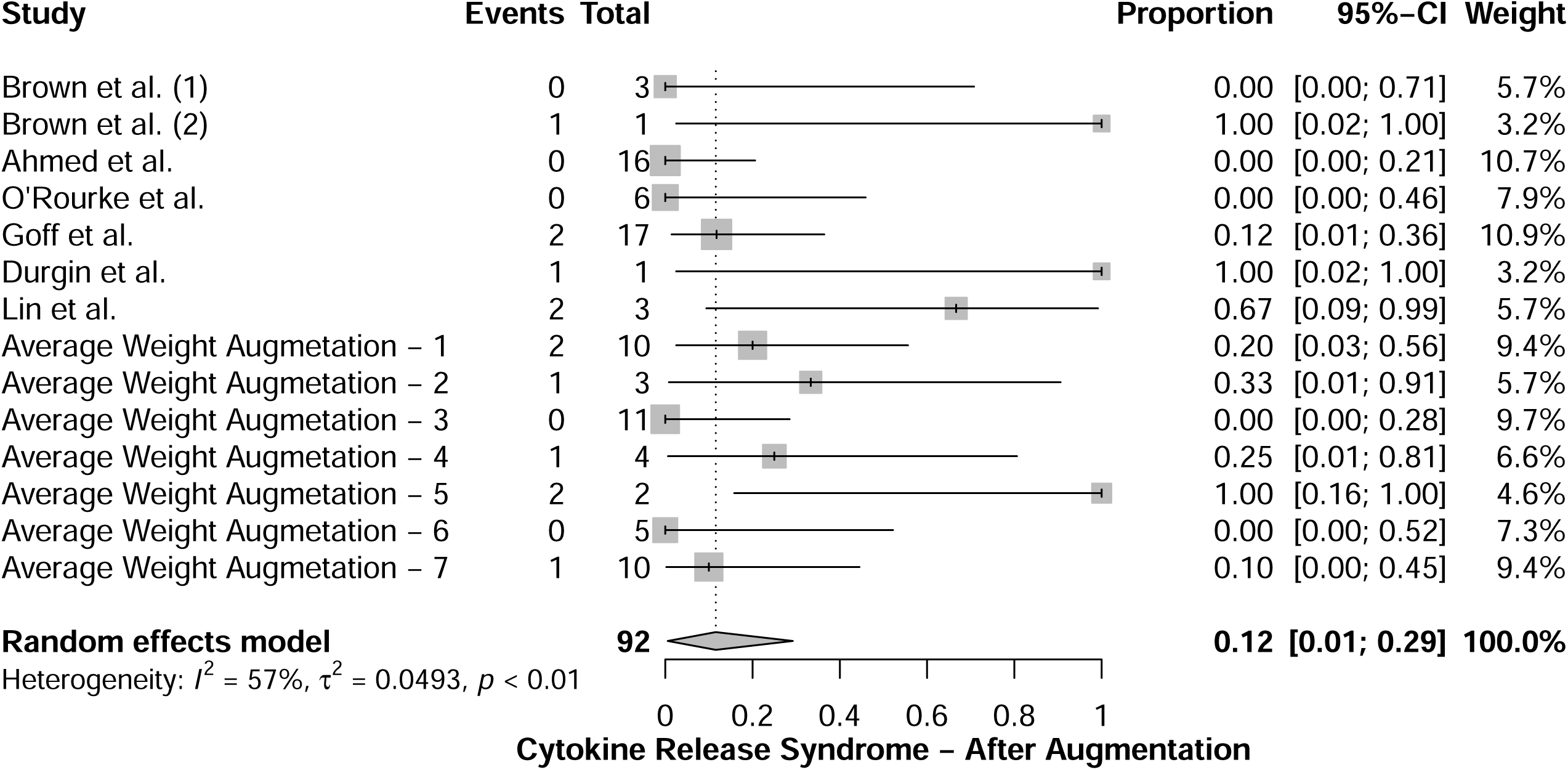
Forest plot for cytokine release syndrome adverse event after augmentation.

Significant heterogeneity was observed (I^2^ = 57%, p-value < 0.01), so the random-effects model was used. Sensitivity analysis was conducted to overcome the significant heterogeneity, after excluding Brown et al. (2), Durgin et al. studies, and the Average Weight Augmentation – 5 the heterogeneity was resolved (I^2^= 29%, p-value= 0.17) resulting in a proportion of **3%, with 95% CI= (0% – 13%)**. Visual inspection of the funnel plot for asymmetry (**Supplementary Figure 12**) did not suggest a risk of publication bias. Egger’s regression test was employed to confirm the presence of publication bias. The results of Egger’s regression test (t = 1.71, df = 9, p-value = 0.1214) clarified that there was no significant risk of publication bias. This signifies a 3% increase in the rate of developing cytokine release syndrome as an adverse event after receiving CAR T-cell therapy compared to the analysis before augmentation.

## 4. Discussion

Our analysis revealed that the mean overall survival for patients with glioblastoma who underwent CAR T-cell therapy was 6.49 months. This outcome did not demonstrate statistical significance compared to the median overall survival observed in patients receiving the current standard protocol for recurrent glioblastoma treatment. Additionally, CAR T-cell therapy did not yield statistically significant results in achieving a complete response within the recurrent glioblastoma cohort. Alarmingly, 58% of these patients experienced worsening of their condition and disease progression. On a more positive note, 44% of patients who received CAR T-cell therapy exhibited a stable disease condition. However, it is crucial to note that CAR T-cell therapy-related encephalopathy was observed in 37% of the treated patients, while only 3% of the patients developed cytokine release syndrome. These findings raise significant concerns regarding the safety profile of CAR T-cell therapy for recurrent glioblastoma patients, as it appears to carry a high risk of adverse events associated with the treatment.

Compared to hematological malignancies, where the objective response rate typically ranges from 57%, and the complete response around 40%, CAR T-cell therapy for glioblastoma patients often results in disease progression [6]. This outcome aligns with findings from prior meta-analyses that primarily focused on solid tumors [6, 7]. We postulate that several factors contribute to this less favorable response. Firstly, unlike hematological malignancies that often feature CD19 as a suitable target with high and stable expression, glioblastoma lacks an ideal target with similar characteristics. For instance, in four studies targeting EGFRvIII, EGFRvIII expression was detected in only 20% of all glioblastoma cases [12, 13]. While IL13Ra2 is expressed in a substantial 80% of glioblastomas, its overexpression is observed in a more limited range, around 50% of glioblastomas [14]. HER2, another potential target, is overexpressed in roughly 10% to 15% of glioblastomas, making it suboptimal due to its lower homogeneity [15].

A promising approach to circumvent this challenge involves using CAR T-cells that target multiple tumor-specific antigens simultaneously. For instance, bi-specific CAR T-cells designed to target both HER2 and IL13Rα2 have demonstrated enhanced tumor elimination in murine glioblastoma models compared to CAR T-cells targeting a single antigen [16]. Taking this concept further, tri-specific CAR T-cells targeting HER2, IL13Ra2, and EphA2 provide broader antigen coverage, with studies indicating that this strategy prolongs the survival of mice bearing glioblastoma patient-derived xenografts [17]. Another obstacle faced during CAR T-cell therapy is the immunosuppressive microenvironment it generates. Combining CAR T-cell therapy with immune checkpoint inhibitors emerges as a promising strategy. While initial clinical trials with immune checkpoint inhibitors as monotherapy did not yield substantial survival benefits, the addition of PD-1/PD-L1 and CTLA4 inhibitors has been found to enhance CAR T-cell activity in murine glioblastoma models [18]. This combination approach holds promise in overcoming the challenges posed by the immunosuppressive milieu within glioblastoma.

Complete responses were exclusively observed in patients who received intracranial or intraventricular administrations of CAR T-cells targeting IL13Ra2. Notably, no complete responses were achieved in patients who underwent intravenous peripheral injections of CAR T-cells targeting other antigens. This observation sheds light on the critical role of the route of administration in CAR T-cell therapy outcomes. Traditionally, it was believed that glioblastomas could breach the blood brain barrier, facilitating the delivery of therapeutic agents to the brain.

However, recent research challenges this notion, suggesting that the blood–brain barrier remains intact even in the presence of glioblastomas with substantial tumor burdens [19]. This intact barrier may impede the successful trafficking of CAR T-cells to intracranial tumors following intravenous administration, potentially explaining the lack of complete responses in this context. In a study involving ten glioblastoma patients treated with intravenous EGFRvIII-specific CAR T-cells, heterogeneous CAR T-cell infiltration was observed in the resected tumor specimens of seven patients who had undergone neurosurgical interventions during the CAR T-cell therapy course [20]. This observation underscores the challenges associated with intravenous delivery and highlights the need for alternative strategies.

Emerging evidence from ongoing clinical trials, such as NCT04045847 and NCT03283631, targeting different antigens (CD147 and EGFRvIII), suggests that intracranial or intraventricular may offer a more effective drug delivery approach in terms of CAR T-cell trafficking. These trials hold promise for improving the efficacy of CAR T-cell therapy by optimizing the route of administration, thereby enhancing the chances of achieving complete responses in glioblastoma patients. Compared to the results of Jang JK et al. [10] meta-analysis, the results of our study do not support the safety and efficacy of CAR-T cell therapy for recurrent glioblastoma. Further phase II and phase III trials are needed to confirm this evidence.

We need to address several limitations in our study. Firstly, there is a limitation related to the relatively small number of trials included in our analysis. To tackle this challenge, we implemented a novel augmentation technique. Secondly, our study’s novelty lies in the application of this augmentation technique, which may not be familiar to many researchers. This statistical method is centered on generating an equivalent number of synthetically generated studies to match the original number of included studies. This approach allowed us to increase the overall number of studies in our quantitative analysis. Consequently, it raised the probability of mitigating potential bias and reduced the concentration of studies favoring a particular direction.

## Conclusions

To our knowledge, this is the first study that utilizes this novel statistical technique to predict the outcomes of CAR T-cell therapy for recurrent glioblastoma. The results of this study are predictive rather than confirmatory. The results of our study shed light on the effectiveness and safety profile of CAR T-cell therapy in recurrent glioblastoma patients. While CAR T-cell therapy has shown promise in some other cancer types, particularly hematological malignancies, our findings reveal a more nuanced picture in the context of glioblastoma. The predicted mean overall survival of 6.49 months in patients who received CAR T-cell therapy is not significantly different from the median overall survival observed in recurrent glioblastoma patients treated with the current standard protocol. This suggests that CAR T-cell therapy, at its current stage of development, does not provide a substantial survival advantage for glioblastoma patients.

Furthermore, the inability to achieve a statistically significant complete response in glioblastoma patients is a notable limitation of CAR T-cell therapy. However, it is encouraging that 44% of the treated patients exhibited stable disease, suggesting some degree of disease control. On the other hand, a concerning finding is that 58% of the patients who received CAR T-cell therapy experienced disease progression. Adverse events related to CAR T-cell therapy were observed in a significant proportion of treated patients, with CAR T-cell therapy-related encephalopathy being a notable concern. Although cytokine release syndrome, another adverse event, was predicted in only 3% of patients, it underscores the importance of careful monitoring and management of side effects associated with this therapy. While CAR T-cell therapy holds promise as a potential treatment for glioblastoma, our study highlights the need for further clinical trials and refinement of this approach to improve its efficacy and safety profile.

Glioblastoma remains a challenging disease, and alternative strategies or combination therapies may be necessary to achieve more significant clinical benefits for patients in the future.

## Declarations

### Ethical Approvals

N/A

### Consent for Participation

N/A

### LLM Statement

We have employed an advanced Large Language Model (LLM) to enhance and refine the English-language writing. This process focused solely on improving the text’s clarity and style, without generating or adding any new information to the content.

## Data Availability

N/A

