## Supplementary figures and images for "Safety and Efficacy of Chimeric Antigen Receptor T-cell Therapy for Recurrent Glioblastoma: An Augmented Meta-analysis of Phase 1 Clinical Trials"

### Supplementary Figure 1

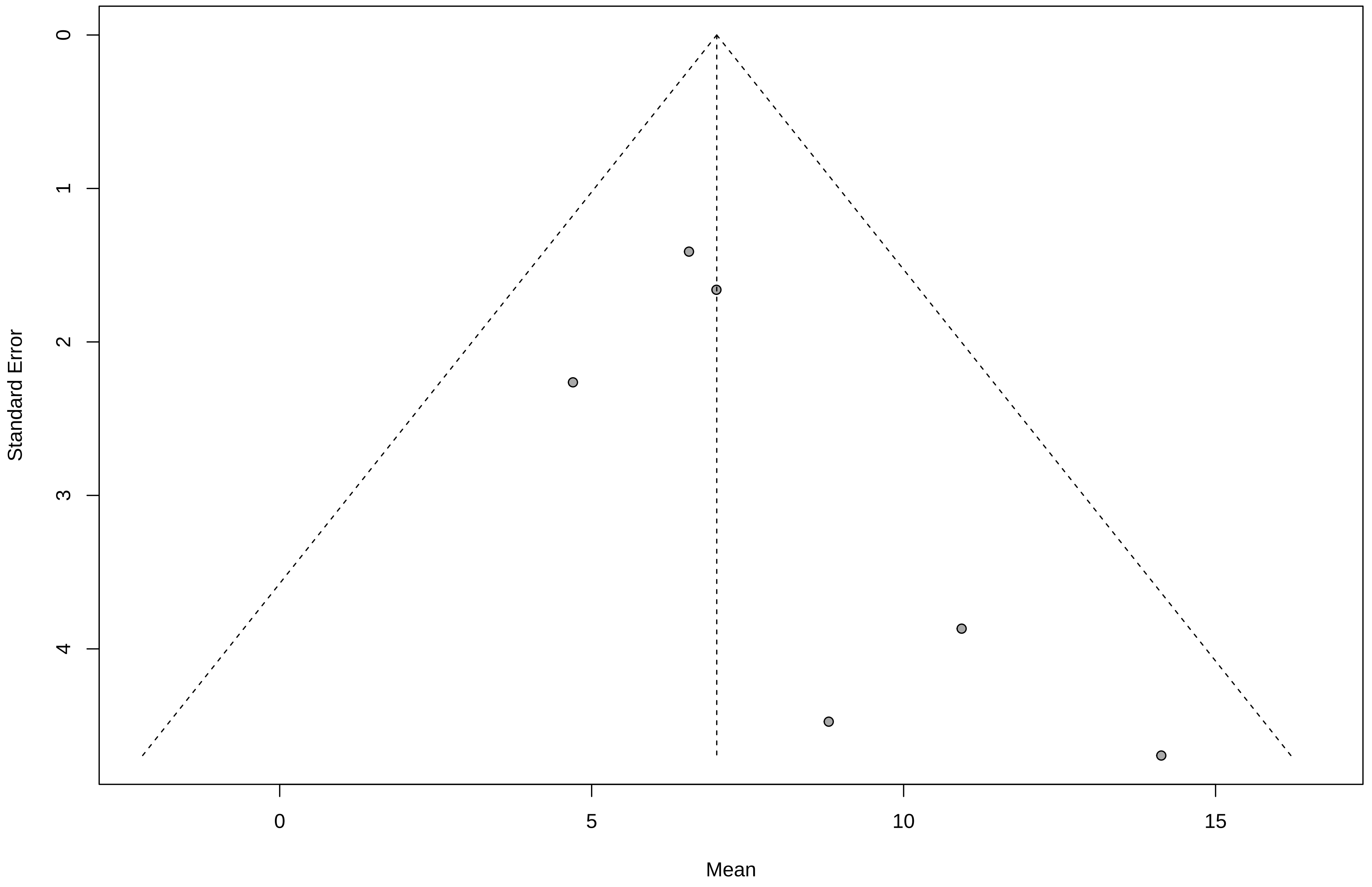

### Supplementary Figure 2

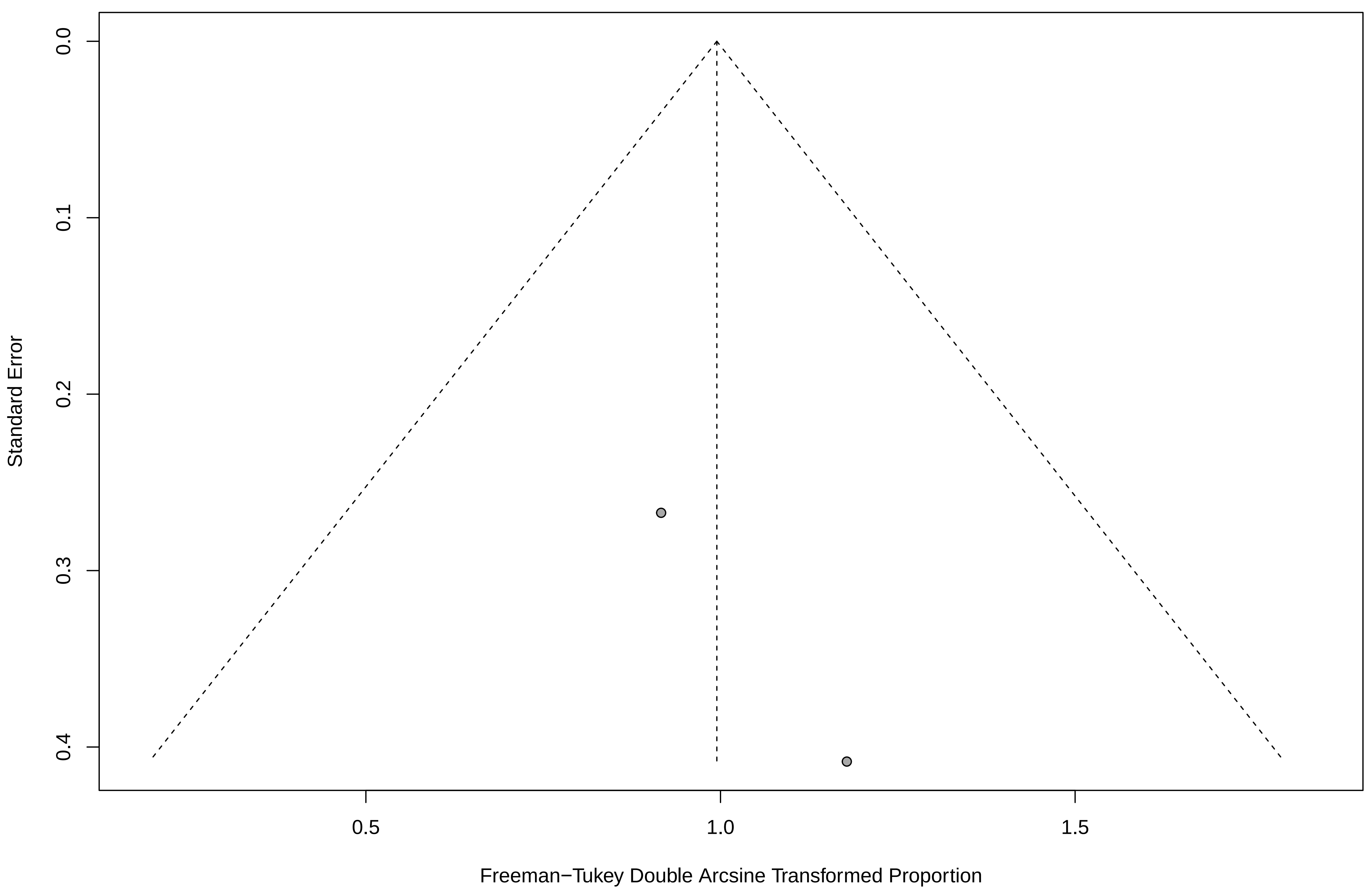

### Supplementary Figure 3

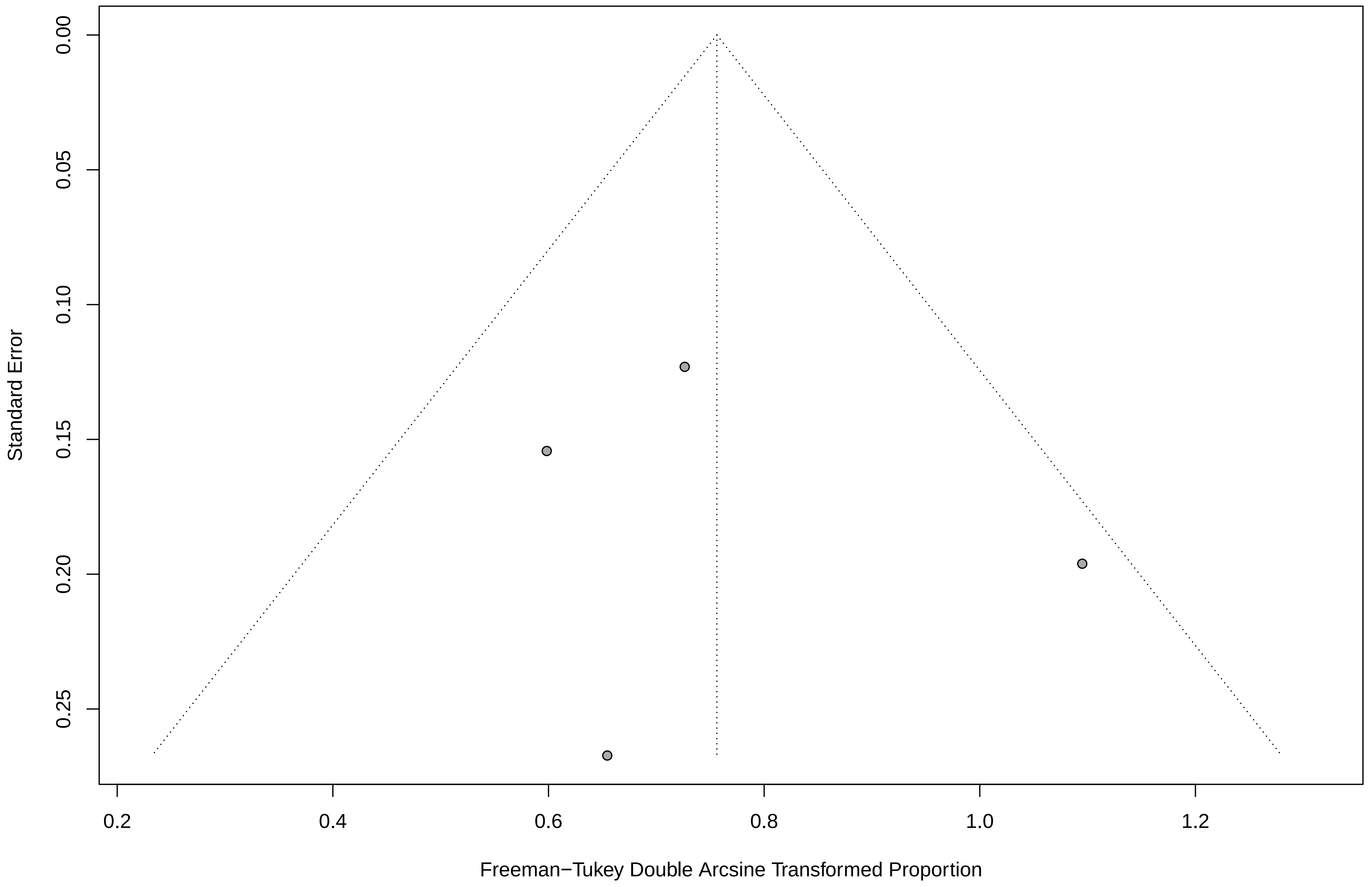

### Supplementary Figure 4

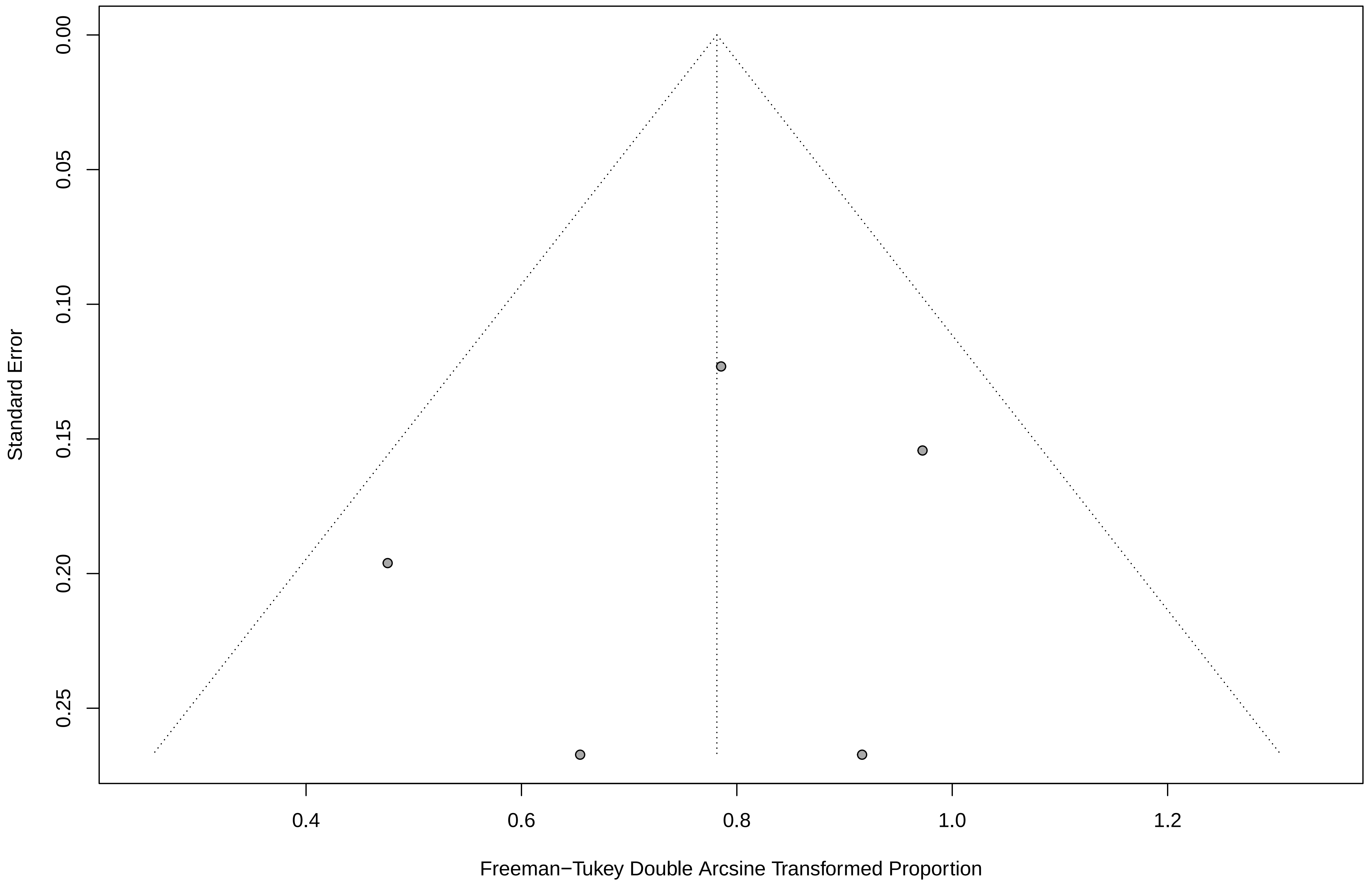

### Supplementary Figure 5

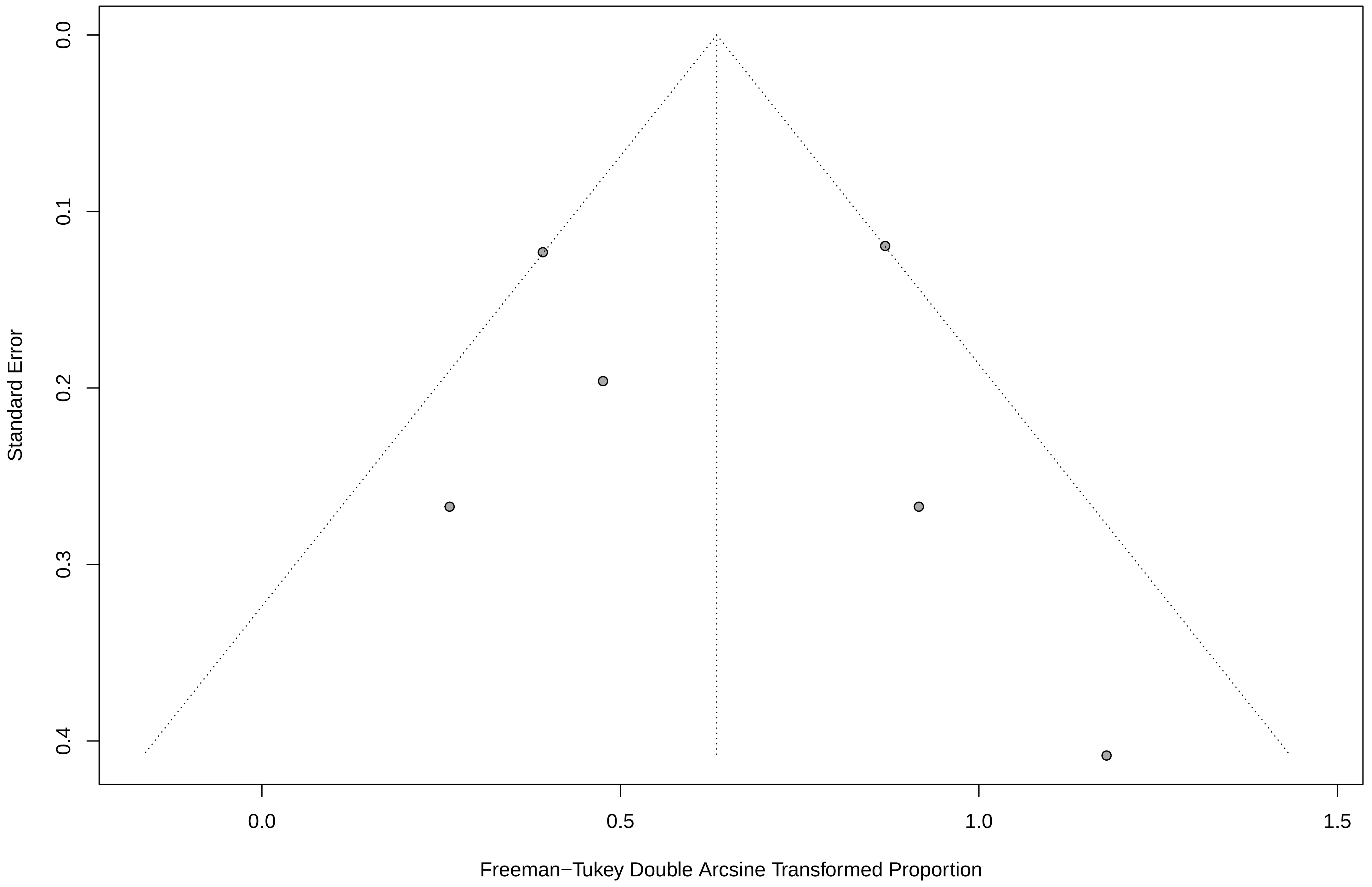

### Supplementary Figure 6

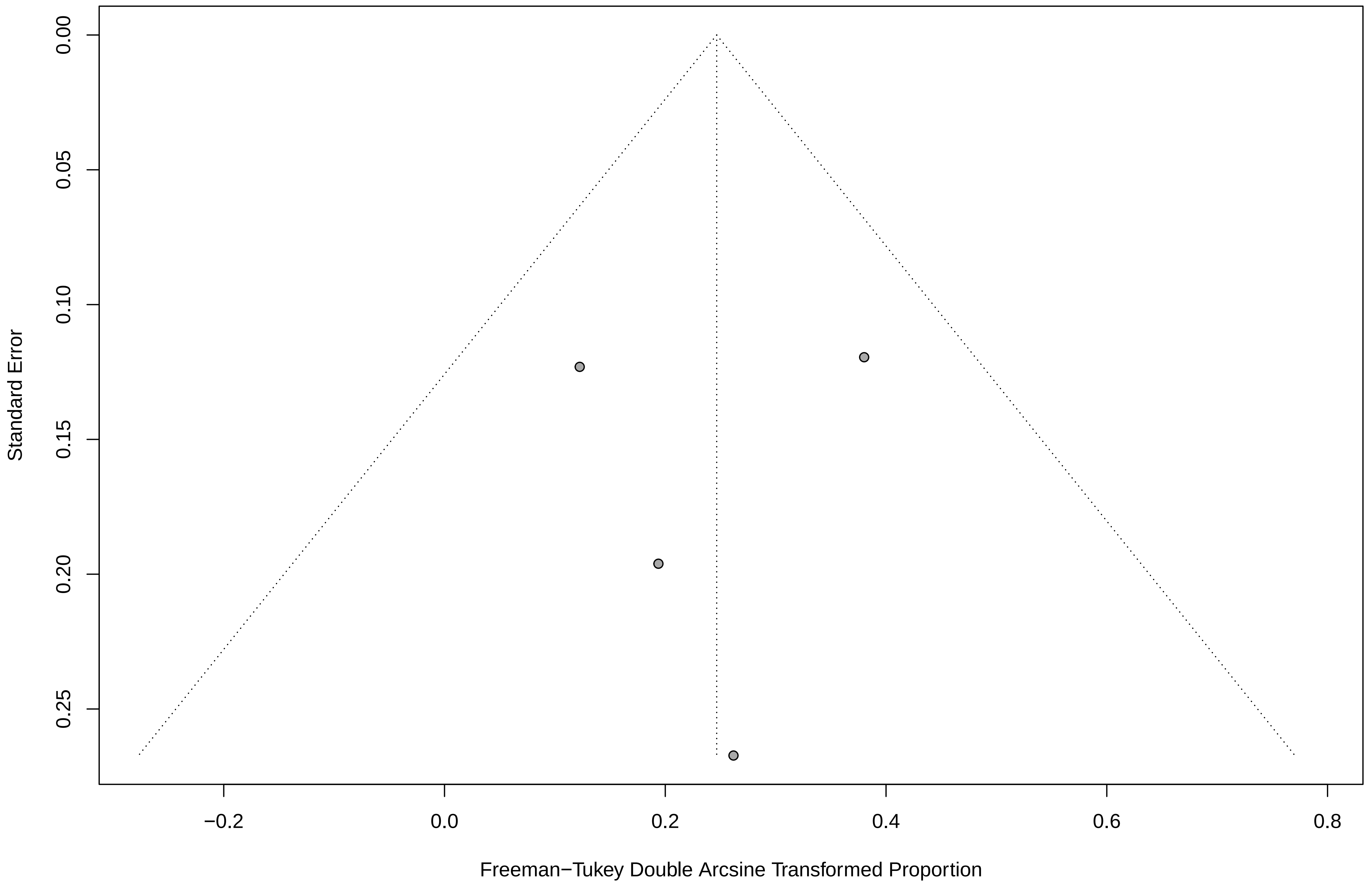

### Supplementary Figure 7

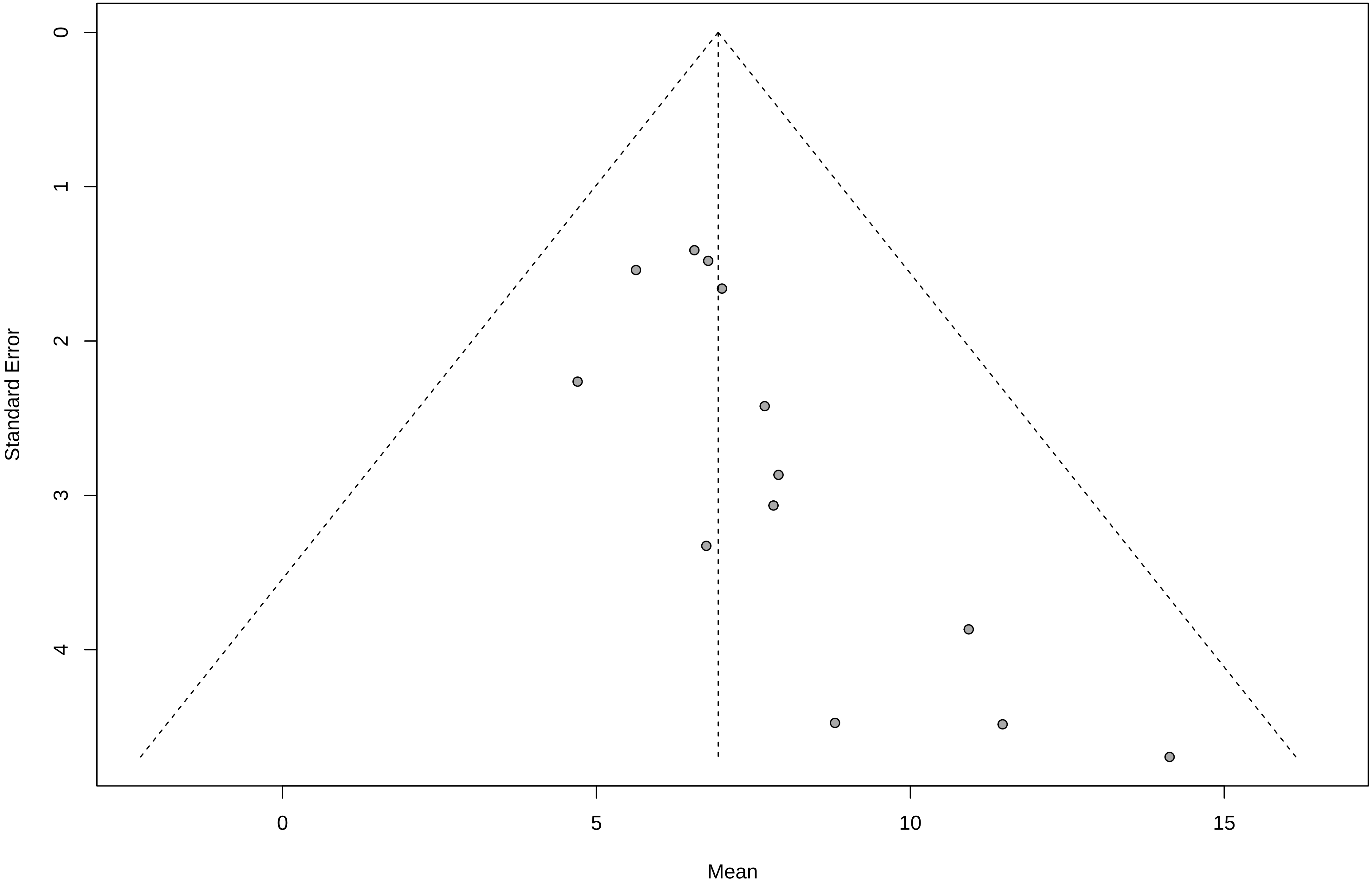

### Supplementary Figure 8

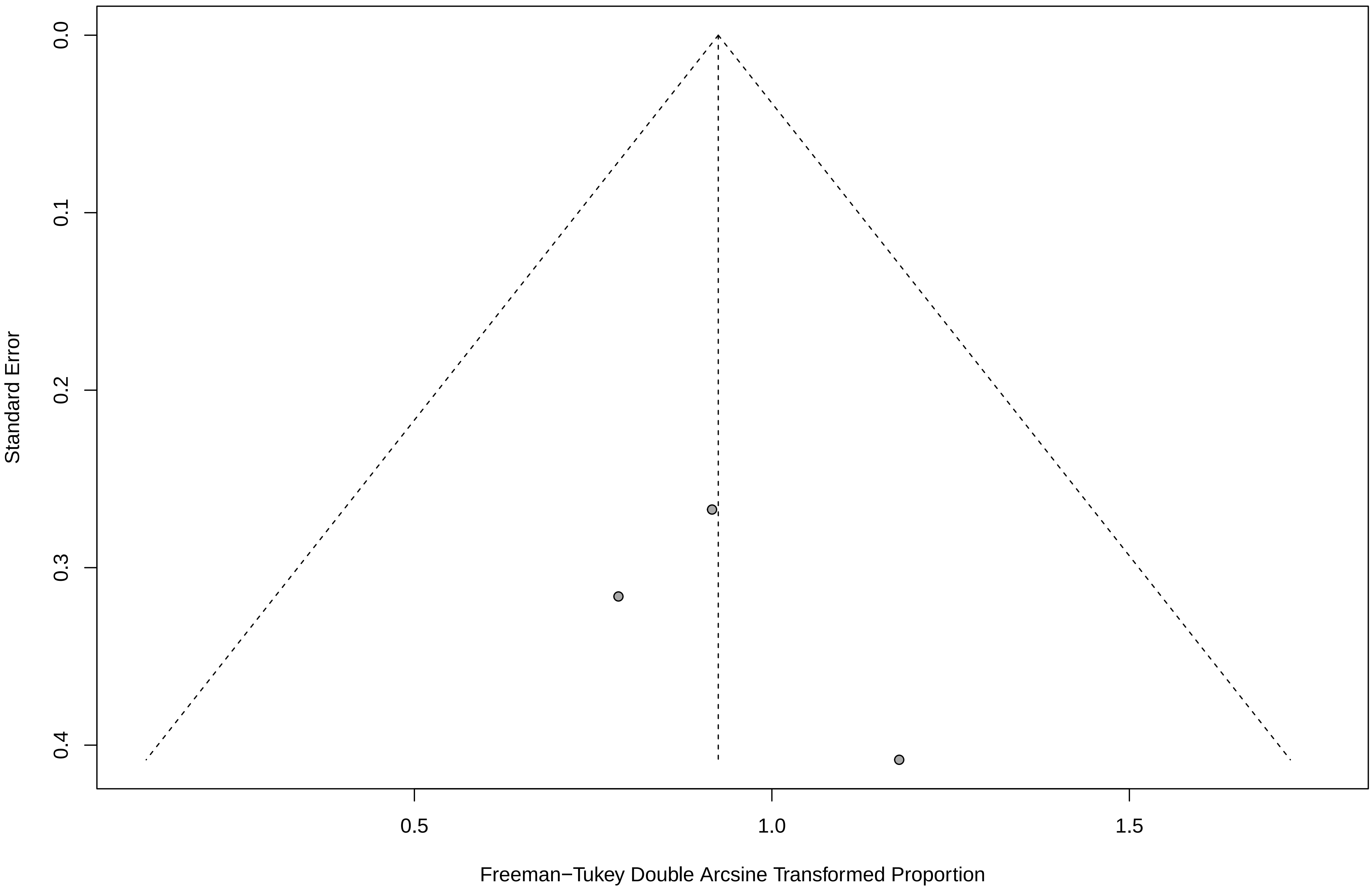

### Supplementary Figure 9

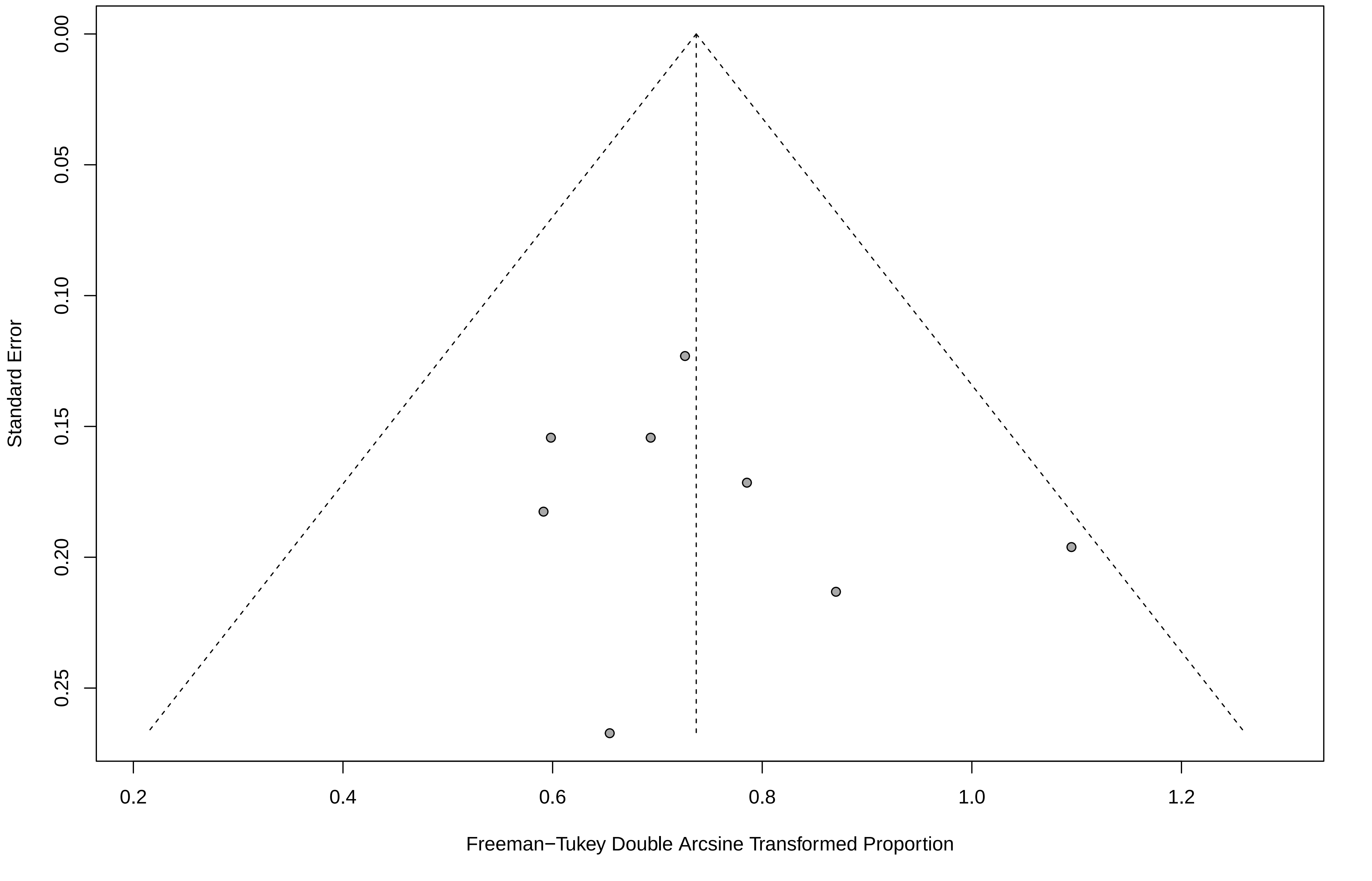

### Supplementary Figure 10

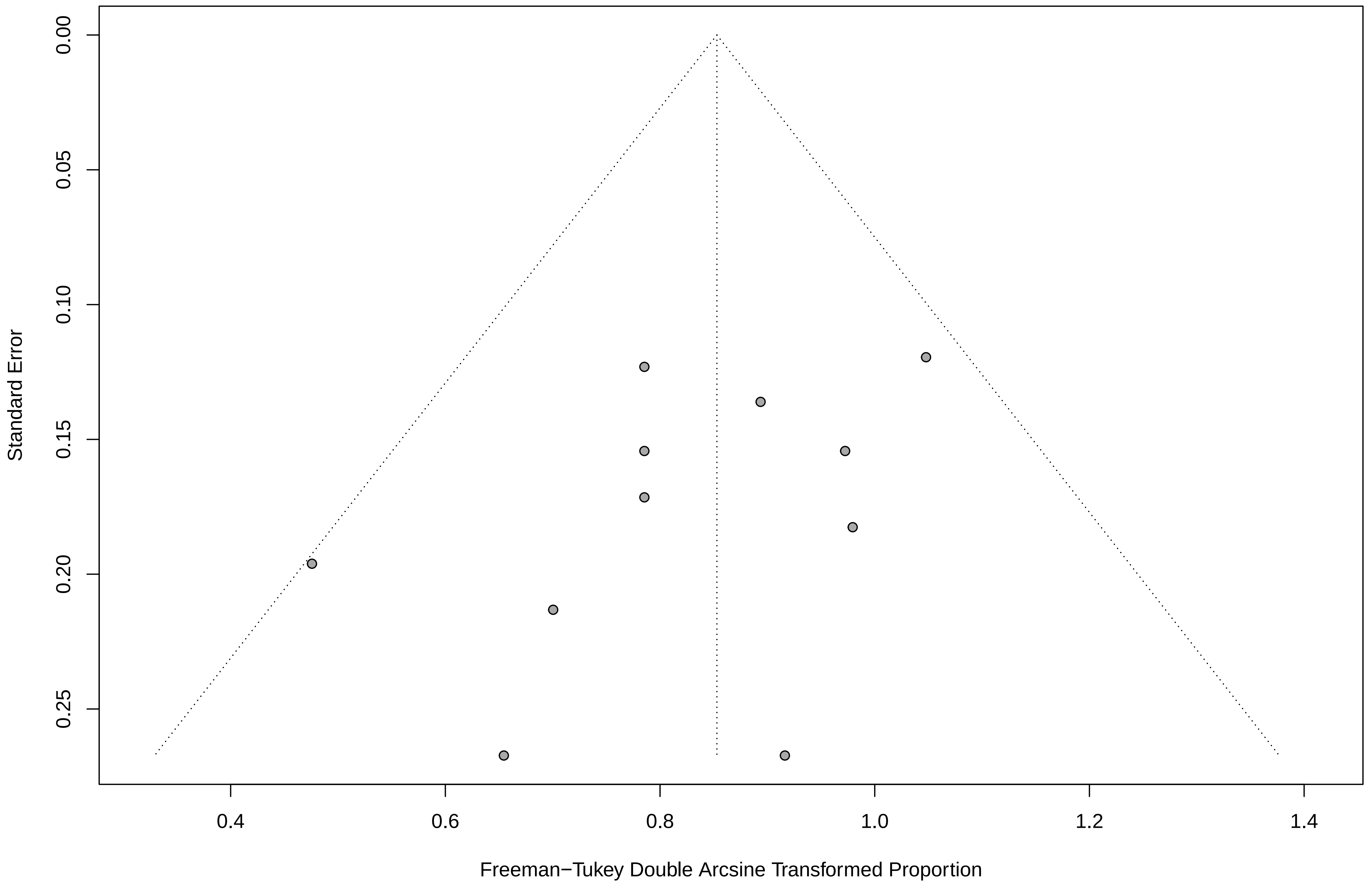

### Supplementary Figure 11

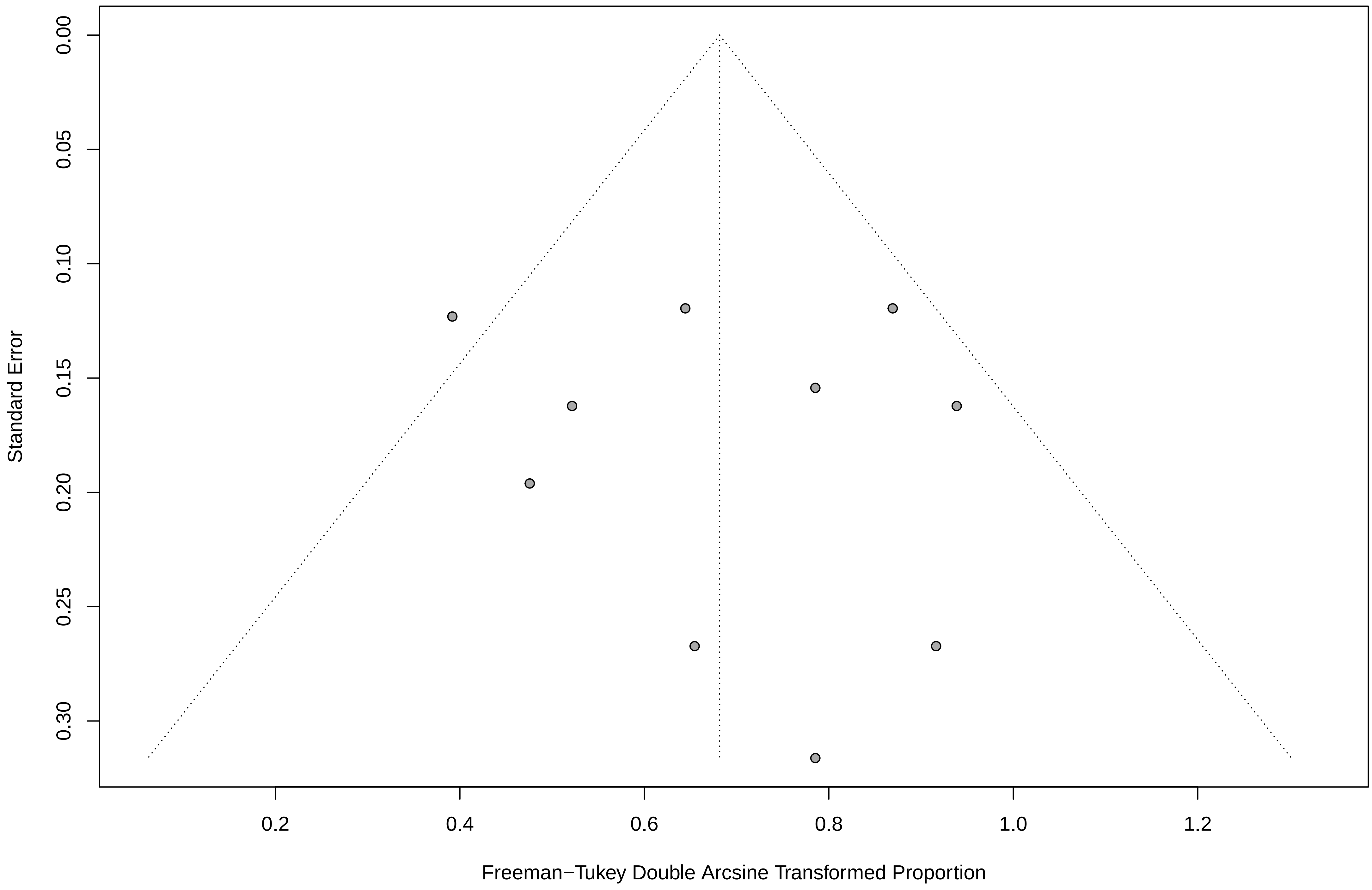

### Supplementary Figure 12

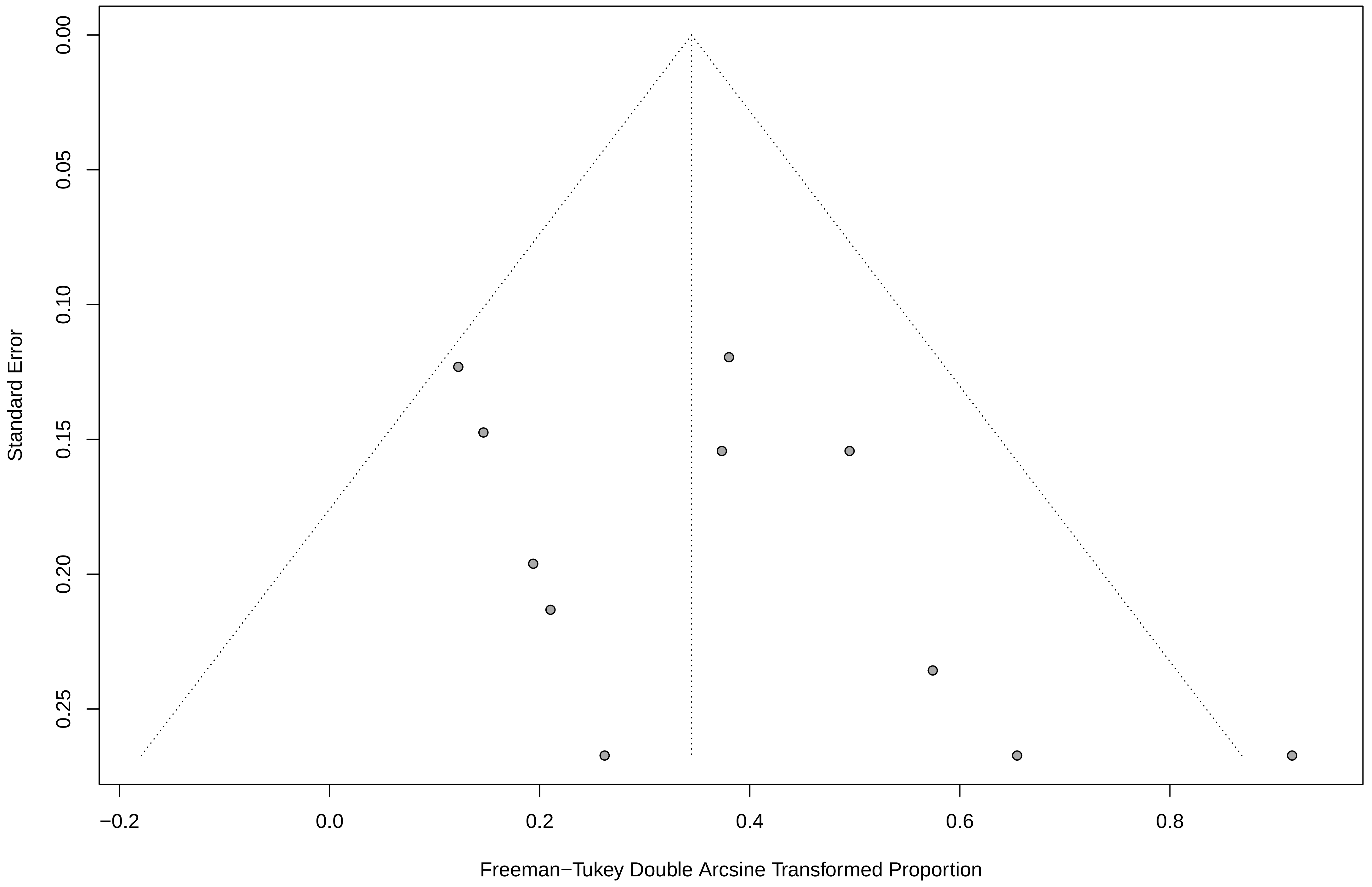
